# Interpretable Lifestyle-Based Machine Learning Models for Ten-Year Cardiovascular Risk Prediction using data from the UK Biobank

**DOI:** 10.64898/2026.01.26.26344438

**Authors:** Yuan Feng, Holger Kunz, Katarzyna Dziopa

**Affiliations:** Institute of Health Informatics, University College London, London, United Kingdom

**Keywords:** Cardiovascular diseases, Risk prediction, Machine Learning, Interpretability, Lifestyle factors, Explainable AI, UK Biobank

## Abstract

**Background:** Cardiovascular diseases (CVDs) remain the leading global cause of morbidity and mortality. In clinical practice, 10-year risk prediction tools such as the Pooled Cohort Equations, QRISK3, and SCORE2 are widely used because of their transparency and clinical trustworthiness, but they rely heavily on biomarkers and medical history. Hence, most recommendations concentrate on pharmaceutical or procedural management, and in many situations, crucial biomarker indicators are unavailable, making it difficult to precisely evaluate individual risk and select appropriate treatments.

**Objective:** To develop interpretable, lifestyle-based machine learning models for predicting 10-year risk of cardiovascular disease (including heart failure and atrial fibrillation), and more critically, to systematically compare interpretability algorithms and assess the cross-model consistency of the identified behavioural factors

**Methods:** Using UK Biobank data, logistic regression, random forest, and XGBoost models were trained on lifestyle (including sleep, smoking, diet, physical activity and electronic device use) and demographic variables only. Discrimination, calibration and interpretability were evaluated using permutation importance, SHapley Additive Explanations and Local Interpretable Model-agnostic Explanations), with subgroup analyses by sex and age to characterise heterogeneity in model behaviour and feature relevance.

**Results:** The developed models demonstrated good discrimination, with XGBoost performing best (ROC-AUC 0.726 [95% CI 0.720–0.731]; PR-AUC 0.199), closely followed by logistic regression (ROC-AUC 0.721 [95% CI 0.716–0.726]; PR-AUC 0.192), while random forest showed slightly lower performance. Despite this similar performance, interpretability analyses revealed inconsistencies in models’ importance ranking of lifestyle factors. Age, sex, and smoking behaviours consistently emerged as key contributors across all interpretability methods, demonstrating strong cross-model agreement, while other lifestyle factors such as dietary patterns, physical activity, and sleep showed model-dependent variation in their assigned importance. Subgroup analyses further indicated that modifiable behaviours (smoking, diet, sleep) were particularly influential among younger females, whereas cumulative exposures and family history were more dominant drivers in older males.

**Conclusions:** Lifestyle-only interpretable models offer a scalable and low-cost framework for cardiovascular risk assessment and behaviour-focused prevention, without requiring laboratory measurements or clinical testing. By comparing multiple interpretability algorithms across models, this study shows strong cross-method consistency and highlights lifestyle factors whose importance profiles differ from those in traditional biomarker-based calculators. These models can complement existing risk tools by highlighting modifiable behaviours, which is particularly valuable for younger adults. They can also support personalised feedback in digital-health settings to promote behavioural change. Overall, the findings support the development of transparent, behaviour-focused tools that enable accessible and equitable cardiovascular prevention.

## Introduction

### Background

#### Cardiovascular Diseases

Cardiovascular diseases (CVDs) are the leading cause of global mortality, responsible for millions of deaths each year and imposing major public health and economic burdens worldwide^1,2^. Chronic heart failure (HF) in particular remains a growing challenge, with mortality rates of 10% within 30 days, 22% within 1 year, and 42% within 5 years after hospital discharge^3^. Although advances in diagnostics and therapeutics have improved outcomes, early and accurate identification of high-risk individuals remains difficult. Reliable risk stratification is essential for prevention, patient management, and optimal allocation of healthcare resources. Notably, more than half of young adults have a low 10-year but high lifetime CVD risk; therefore, regular screening and early behavioural interventions can substantially reduce disease onset ^4^. Consequently, multiple risk prediction tools have been developed to support preventive strategies and reduce both mortality and healthcare costs.

Cardiovascular risk prediction tools have long supported prevention efforts in primary care. Current guideline-recommended models, including SCORE2^5,6^ in Europe, QRISK3^7^ in the UK, and the Pooled Cohort Equations^8^ in the US, are all based on transparent Cox proportional hazards frameworks that provide clear, interpretable associations between predictors and outcomes. These tools typically rely on routinely collected clinical predictors and medication history, many of which require laboratory measurements or clinical assessments. This transparency enables clinicians to validate model behaviour, enhance clinical and patient trust, understand key risk drivers, and effectively communicate results during consultations. Their widespread adoption underscores a broader principle: interpretability is often essential for clinical acceptance, shared decision-making, and real-world implementation.

#### Artificial Intelligence and Machine Learning in CVD

Artificial intelligence (AI) and machine learning (ML) including deep neural networks, gradient boosting models, and other complex non-linear algorithms, have demonstrated strong predictive performance in cardiovascular research^9^, but remain underused in clinical practice^10^. Many high-performing “black-box” models provide little insight into how predictions are generated, which reduces clinical trust and hinders external validation^11^. In addition, inconsistent calibration and limited generalizability across populations further constrain their real-world applicability. In addition, inconsistent calibration and limited generalizability across populations further constrain their real-world applicability.

Interpretability is therefore critical for detecting bias, improving clinician confidence, and supporting shared decision-making, while explainability techniques such as SHapley Additive Explanations (SHAP)^12^ and Local Interpretable Model-agnostic Explanations (LIME)^13^ enhance transparency by showing how individual features contribute to predicted risk and by revealing potential model flaws. These examples highlight the need for new approaches that combine predictive accuracy with transparency and usability, providing interpretable and trustworthy decision support for cardiovascular risk prediction.

#### Lifestyle Factors in CVDs

Lifestyle predictors are selected because they are modifiable, easily measurable, and directly linked to behavioural interventions, enabling early, low-cost risk assessment. A comprehensive meta-analysis of 22 prospective cohort studies reported that adherence to multiple healthy behaviours was associated with a 66% reduction in cardiovascular disease (CVD) risk, a 69% reduction in heart failure, and a 60% reduction in stroke risk compared with adopting none or only one healthy behaviour, showing a clear dose–response relationship^14^. Consistent findings from large-scale cohort and meta-analytic studies further confirm that maintaining multiple healthy behaviours substantially lowers the risks of CVD, mortality, and multimorbidity across chronic conditions^15^. Moreover, recent analyses from UK Biobank and National Health and Nutrition Examination Survey (NHANES) indicate that accelerated biological aging partly mediates these associations^16^, reinforcing the biological and preventive rationale for developing lifestyle-based, interpretable prediction models to reduce risk through reasonable and well-planned lifestyle regulations.

#### Aim

This study aims to develop and evaluate clinically interpretable machine learning models for predicting 10-year risks of CVD, HF, and atrial fibrillation (AF) using UK Biobank data. The models with clinical acceptance performance integrate interpretability and transparency to enhance clinical relevance, trust, and communication. Predictor variables are drawn from lifestyle domains (smoking, alcohol intake, physical activity, diet, sleep, and psychosocial factors) and basic demographic characteristics (age and sex), identified in prior work by Dziopa et al^17,18^. Three algorithms, logistic regression (LR), random forest (RF), and XGBoost, are implemented and compared in terms of discrimination, calibration, and explainability using SHAP^12^for global explanations and LIME^13^ for local explanations, while subgroup analyses by age and sex examine the consistency and fairness of predictions and feature attributions.

The study addresses two key questions: (1) which lifestyle factors most strongly influence 10-year risks of CVD, HF, and AF; and (2) how predictive performance, model behaviour, interpretability, and feature importance differ across LR, RF, and XGBoost models, with these contrasts further clarified using complementary interpretability algorithms that provide detailed insight into model reasoning, together with subgroup analyses by sex and age to examine heterogeneity.

## Methods

### Recruitment

#### Study Design and Data Source

This study used data from the UK Biobank^19^, a large prospective cohort of over 500 000 adults aged 40–69 years at baseline, containing extensive phenotypic, lifestyle, and linked health records. To ensure clinical relevance and minimise reverse causality, individuals with a history of cardiovascular disease (CVD), heart failure (HF), or atrial fibrillation (AF) at baseline were excluded based on hospital and self-reported data.

Model development focused exclusively on lifestyle-related predictors, deliberately omitting clinical biomarkers; these indicators, although associated with CVD in previous research, were not used for model training but rather to characterise the study cohort and emphasise modifiable behavioural risk factors. Selected clinical indicators were described solely to contextualise baseline health status rather than to inform model training.

#### Patient Level Clinical Baseline

At baseline, clinical characteristics of the study population were summarised to provide an overview of the cohort and to contextualise subsequent analyses (see Table 1). The study included 459,142 participants with a mean age of 56.2 years (SD 8.1). The median BMI was 26.5 kg/m^2^ (IQR 24.0–29.6), while mean systolic and diastolic blood pressures were 139.6 mmHg and 82.3 mmHg, respectively. Lipid profiles showed average HDL, LDL, and total cholesterol levels of 1.47, 3.62, and 5.79 mmol/L. Inflammatory and metabolic markers were also captured, with a mean CRP of 2.5 mg/L, HbA1c of 35.3 mmol/mol, and creatinine of 71.7 µmol/L. Females accounted for 43.9% of the cohort (n = 201,782) and males for 56.1% (n = 257,360). During follow-up, 8.8% of participants (n = 40,350) experienced the composite outcome of CVD, HF and AF.

**Table 1.**
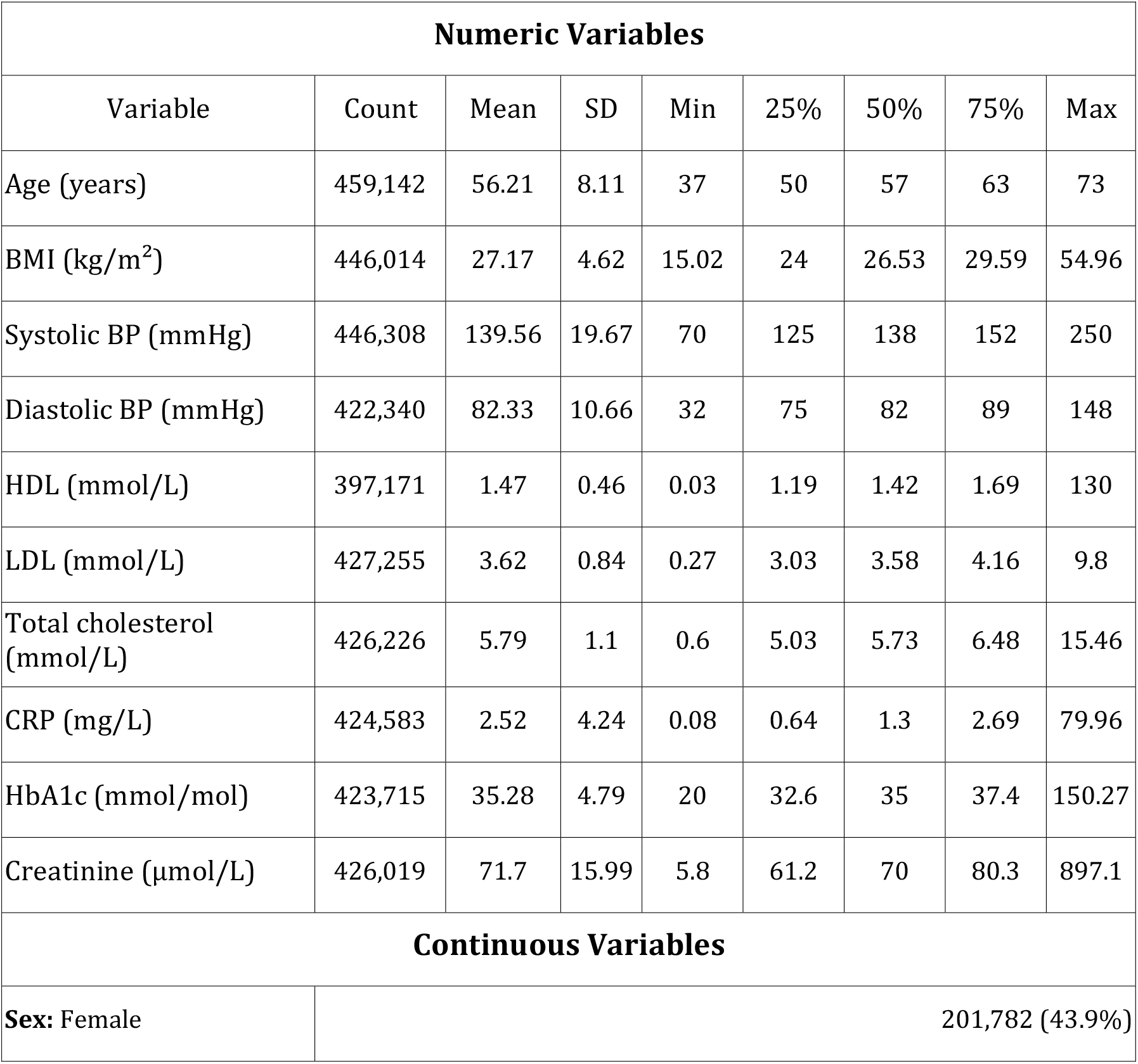
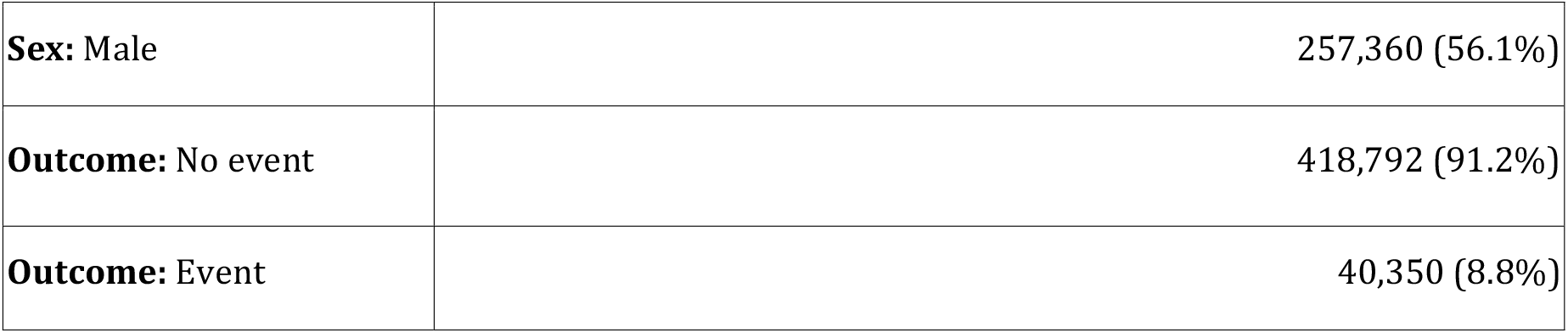
Baseline characteristics of the study population (N = 459142). Continuous variables are summarised with count, mean, standard deviation, and quantiles; binary variables are shown as N%.

| Numeric Variables |  |  |  |  |  |  |  |  |
| --- | --- | --- | --- | --- | --- | --- | --- | --- |
| Variable | Count | Mean | SD | Min | 25% | 50% | 75% | Max |
| Age (years) | 459,142 | 56.21 | 8.11 | 37 | 50 | 57 | 63 | 73 |
| BMI (kg/m<sup>2</sup>) | 446,014 | 27.17 | 4.62 | 15.02 | 24 | 26.53 | 29.59 | 54.96 |
| Systolic BP (mmHg) | 446,308 | 139.56 | 19.67 | 70 | 125 | 138 | 152 | 250 |
| Diastolic BP (mmHg) | 422,340 | 82.33 | 10.66 | 32 | 75 | 82 | 89 | 148 |
| HDL (mmol/L) | 397,171 | 1.47 | 0.46 | 0.03 | 1.19 | 1.42 | 1.69 | 130 |
| LDL (mmol/L) | 427,255 | 3.62 | 0.84 | 0.27 | 3.03 | 3.58 | 4.16 | 9.8 |
| Total cholesterol (mmol/L) | 426,226 | 5.79 | 1.1 | 0.6 | 5.03 | 5.73 | 6.48 | 15.46 |
| CRP (mg/L) | 424,583 | 2.52 | 4.24 | 0.08 | 0.64 | 1.3 | 2.69 | 79.96 |
| HbA1c (mmol/mol) | 423,715 | 35.28 | 4.79 | 20 | 32.6 | 35 | 37.4 | 150.27 |
| Creatinine (µmol/L) | 426,019 | 71.7 | 15.99 | 5.8 | 61.2 | 70 | 80.3 | 897.1 |
| Continuous Variables |  |  |  |  |  |  |  |  |
| Sex: Female | 201,782 (43.9%) |  |  |  |  |  |  |  |

|  |  |
| --- | --- |
| **Sex:** Male | 257,360 (56.1%) |
| **Outcome:** No event | 418,792 (91.2%) |
| **Outcome:** Event | 40,350 (8.8%) |

#### Predictor Variables and Feature Processing

A total of 79 lifestyle-relevant features were selected based on clinical relevance and data availability. When mapped to their corresponding UK Biobank field IDs, several multi-category variables expanded into multiple columns, resulting in 91 derived variables after preprocessing. After adding age and sex for subgroup stratification, the final analytic dataset comprised 93 variables. Variables were selected for clinical relevance and data availability following the curation strategy described by Dziopa et al^17,18^

Multi-category variables were one-hot encoded to enhance interpretability. Missing values were encoded using a unique constant value to preserve meaningful structural missingness, particularly for smoking-related variables where unanswered items occur only among non-smokers. Retaining these coded entries minimises selection bias and allows the models to capture informative missingness patterns without fabricating behavioural values, reflecting cases where missingness itself carries behavioural information.

The final set of 79 lifestyle features covered diet, physical activity, sleep, alcohol and tobacco use, and psychosocial domains. The dataset was randomly divided into training and testing sets (80:20) using fixed participant identifiers and a random seed to ensure reproducibility and prevent data leakage. The binary outcome variable was coded as 1 (event) and 0 (non-event) to ensure consistency with standard binary classification input formats.

### Statistical Analysis

#### Model Development and Hyperparameter Tuning

Three supervised learning models were implemented using the pre-defined training and test subsets: Logistic Regression^20^, Random Forest^21^, and XGBoost^22^ to balance interpretability and predictive performance. LR provided a transparent linear baseline; RF captured non-linear relationships through ensemble averaging, and XGBoost offered a high-performing gradient boosting framework suitable for structured data. Class imbalance was addressed using model-specific weighting schemes, and feature standardisation was applied only to LR. All models used the same predefined train–test split to ensure fair comparison.

Hyperparameters were tuned over predefined ranges for each algorithm using stratified cross-validation, and the combination that achieved the highest mean ROC-AUC was selected. Full technical specifications, including the hyperparameter search spaces, cross-validation settings, and the threshold ranges tested to examine their impact on predictive performanc. After tuning, operating thresholds were evaluated to support different decision-making scenarios, and final models were selected based on test-set performance. Model uncertainty was quantified using bootstrap-derived confidence intervals.

#### Model Evaluation

Model performance was evaluated using complementary discrimination and calibration metrics to capture both predictive accuracy and the reliability of estimated risks. Discrimination was primarily assessed using the C-statistic (ROC-AUC), indicating the model’s ability to rank individuals correctly. To reflect performance at clinically relevant thresholds, additional measures including sensitivity, specificity, precision, and F1-score were examined. Calibration performance was assessed using Brier scores and calibration curves, with probabilistic recalibration applied through Platt scaling to evaluate how closely predicted probabilities matched observed event rates. Together, these complementary metrics provide a comprehensive view of overall model.Model Explainability and Robustness

To ensure transparency and clinical interpretability^23^, complementary global and local explanation methods were applied. Global feature importance was quantified using both model-specific and model-agnostic approaches: impurity-based importance for tree models and permutation importance^24^ for LR to account for coefficient shrinkage under regularisation. Model-agnostic SHAP^12^ across all algorithms to capture each feature’s marginal contribution. Mean absolute SHAP values on the test set summarised overall influence, while summary and dependence plots visualised effect direction. LIME^13^ provided case-specific explanations using sparse linear approximations on the top 20 SHAP-ranked features, balanced between event and non-event cases. To link interpretability to performance, permutation-based changes in the C-statistic were calculated to assess each feature’s impact on discrimination. Together, these methods captured both marginal and global influences, enabling consistent interpretation of feature relevance and providing clinicians with an understanding of how lifestyle factors shape individual and population-level risk patterns.

Fairness and generalisability were assessed through subgroup analyses stratified by sex and age quartiles. The cohort was divided by sex (female, male) and age quartiles (Q1–Q4), yielding eight demographic strata. Evidence indicates that AI-based cardiovascular prediction may vary by sex and age due to data imbalance or structural bias^25^. Subgroups with sufficient sample size (n ≥ 500) were evaluated using the same trained models to isolate genuine performance differences rather than retraining effects. SHAP values were derived from the pooled test set and partitioned by subgroups to maintain a consistent reference scale. Mean absolute SHAP values and ranked feature importances were then compared to assess the stability of predictive accuracy and dominant lifestyle contributors across strata, enabling detection of demographic heterogeneity and supporting equitable, subgroup-informed risk evaluation.

#### Ethical Considerations

This research has been conducted using the UK Biobank Resource under application numbers 12113. We are grateful to the UK Biobank participants. UK Biobank was established by the Wellcome Trust medical charity, Medical Research Council, Department of Health, Scottish Government, and the Northwest Regional Development Agency. The source code supporting the findings of this study is available at https://gitlab.com/Iris9F/interpretable-lifestyle-cvd-risk.

## Results

### Model Performance

Three supervised machine-learning models (LR, RF, and XGBoost) were developed and evaluated. Table 2 summarises discrimination and calibration performance across models. XGBoost achieved the highest discrimination (ROC-AUC = 0.752, 95 % CI [0.747–0.757]; PR-AUC = 0.230), followed closely by Logistic Regression (ROC-AUC = 0.748, 95 % CI [0.743–0.753]; PR-AUC = 0.225). RF showed weaker calibration. SMOTE oversampling did not improve recall, indicating limited utility for structured epidemiological data. Hence, LR and XGBoost were retained for interpretability and subgroup analyses.

**Table 2.** Overall calibration and discrimination performance across three models.

| Model | PR-AUC (Test) | ROC-AUC (Test) | 95% CI (ROC-AUC) |
| --- | --- | --- | --- |
| Logistic Regression | 0.192 | 0.721 | [0.716 – 0.726] |
| Random Forest | 0.187 | 0.717 | [0.711 – 0.722] |
| **XGBoost** | **0.199** | **0.726** | **[0.720 – 0.731]** |

In the original QRISK3^7^ development study, Harrell’s C was reported as 0.88 in women and 0.86 in men. Similarly, SCORE2^26^ achieved C-indices ranging from 0.67 to 0.81 across European regions. These tools incorporate clinical and laboratory markers, whereas the present study intentionally evaluates lifestyle-only prediction performance. Compared with established clinical risk algorithms, our models show lower discrimination, which is expected given that they rely solely on lifestyle variables without clinical biomarkers.

XGBoost demonstrated the strongest performance among all models. Hyperparameter tuning focused on learning rate, tree depth, and number of trees (with *subsample* and *colsample_bytree* fixed at 0.8), yielding an optimal configuration of a shallow model (learning rate = 0.1, depth = 3, 200 trees). This model achieved a mean cross-validated ROC-AUC of 0.728 ± 0.0018 and a test ROC-AUC of 0.726 (95% CI 0.720–0.731), corresponding to a PR-AUC of 0.199. In the overall comparison (*see Table 1*), XGBoost achieved the highest discrimination (ROC-AUC = 0.752; PR-AUC = 0.230). The ROC and precision–recall curves (*see Appendix section 5*) showed a steeper rise and consistently higher precision across recall levels compared with other models, indicating superior ranking and probability calibration. Threshold analysis identified an F1-optimal cut-off of 0.55 (F1 = 0.265, Recall = 0.607, Precision = 0.170), while the recall-focused “Policy A” threshold of 0.50 achieved Recall = 0.706 and Precision = 0.169, demonstrating a balanced sensitivity–precision trade-off.

For comparison, the logistic regression model tuned the regularisation strength *C* (0.01–2) under L2 penalty using the *lbfgs* solver, achieving stable and interpretable performance (CV ROC-AUC = 0.724 ± 0.0016; test ROC-AUC = 0.721, 95% CI 0.716– 0.726; PR-AUC = 0.192). While logistic regression exhibited well-calibrated probabilities and competitive discrimination (AUC = 0.748; PR-AUC = 0.225 in the full comparison), XGBoost ultimately outperformed it in both discrimination and recall, providing a more flexible and reliable representation of complex lifestyle–risk interactions.

### Calibration

Figure 1 illustrates the calibration performance of the three models after sigmoid calibration, with Brier scores of 0.076 forLR, 0.075 for RF, and 0.074 for XGBoost. All models showed reasonable calibration, but with notable differences: LR aligned well at lower probabilities but diverged above 0.3 due to sparse samples in higher bins, RF exhibited a similar pattern with limited resolution at the upper end, and XGB followed the diagonal more closely, slightly overestimating risk in the mid–high range. Overall, XGB produced the most reliable probability estimates, while the apparent miscalibration of LR and RF at higher probabilities should be interpreted cautiously as a reflection of small sample counts rather than systematic bias.

**Figure 1.**
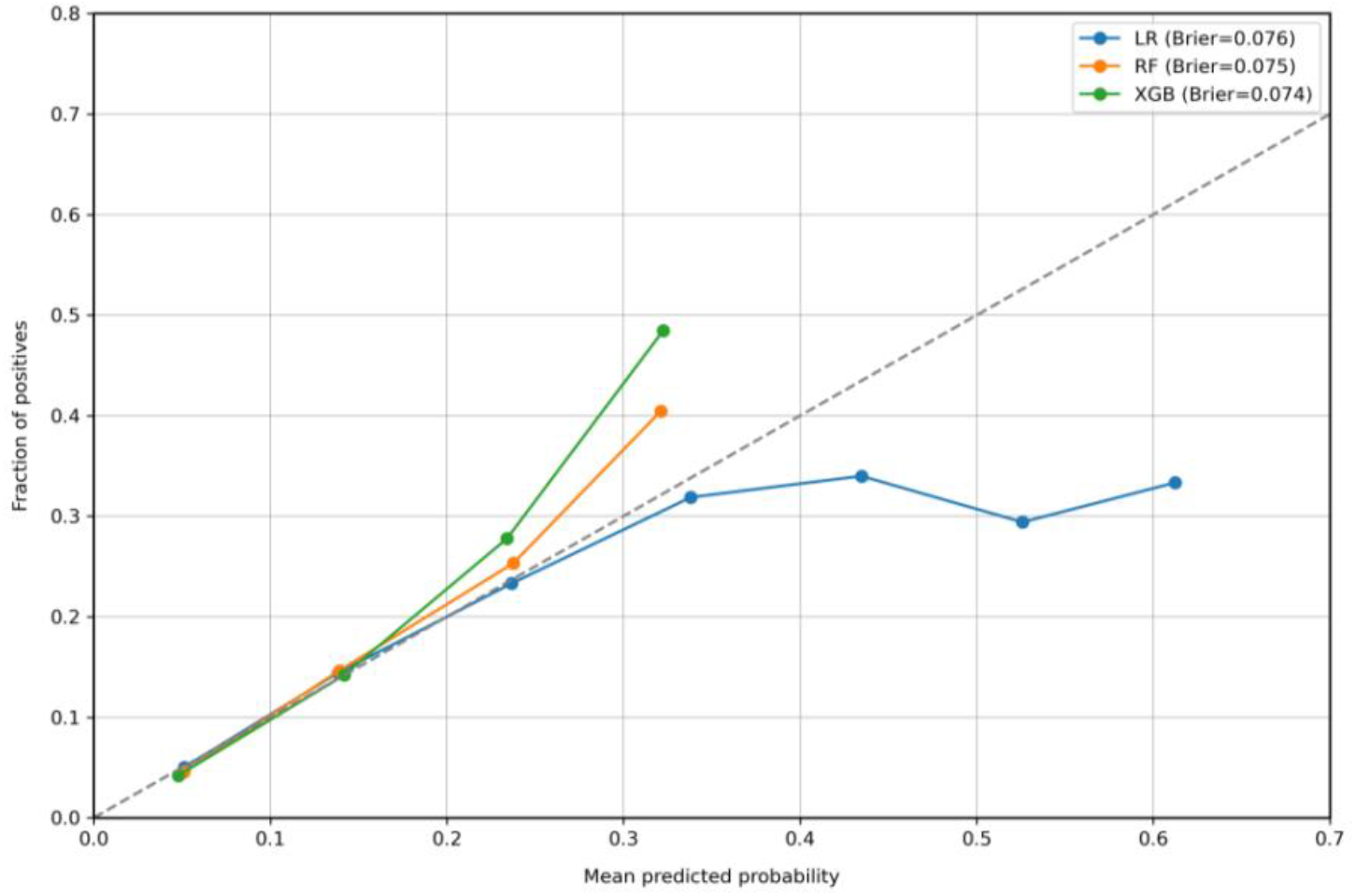
Calibration curves (test set) for LR, RF and XGB.

**Figure 2.**
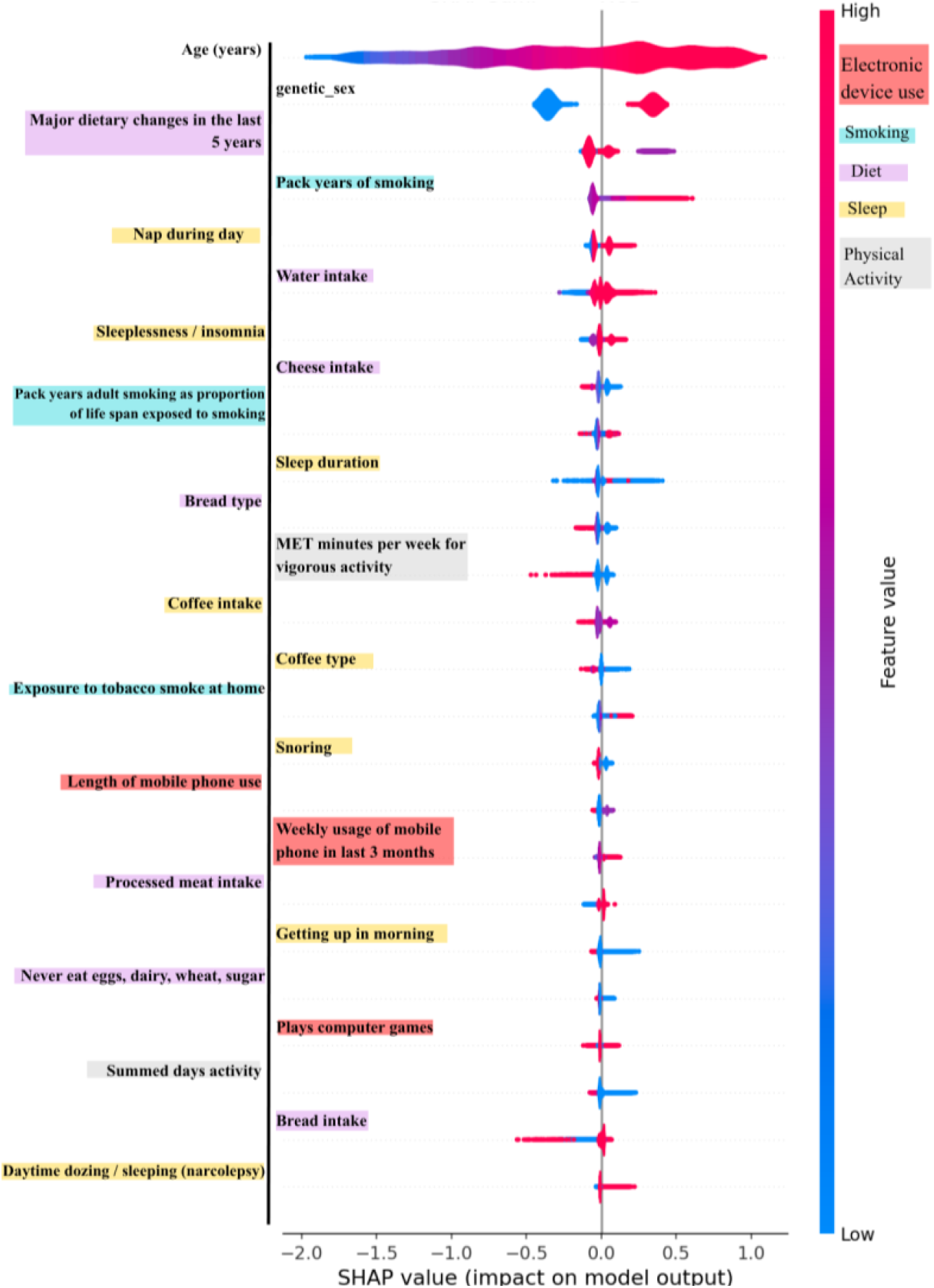
SHAP summary plots for XGBoost. The distribution of SHAP values by feature.

### Feature Importance

#### Model-Derived Importance

Feature importance analyses across models revealed consistent yet nuanced patterns *(see Appendix table 10)*. All models identified age and sex as the strongest predictors, highlighting their foundational and well-established role in shaping long-term cardiovascular risk. Beyond demographics, smoking was the most stable and influential across models, with cumulative exposure, particularly pack-years and smoking intensity, consistently ranking among the highest-impact features.

Tree-based models (RF and XGBoost) captured a broader spectrum of behavioural signals compared with logistic regression, reflecting their capacity to model non-linear interactions among lifestyle domains. In these models, dietary patterns, sleep behaviours, and physical activity metrics collectively demonstrated meaningful but heterogeneous contributions, suggesting that these behaviours influence cardiovascular risk through complex, multi-dimensional pathways rather than simple linear effects. In particular, XGBoost assigned notable weight to sleep-related indicators, whereas RF placed greater emphasis on physical activity MET scores and cumulative activity time. Physical activity measures were also more prominent in ensemble models, indicating that supporting cumulative movement patterns carry more predictive signal.

Overall, the convergence across methods indicates that interpretable ML recovers a coherent structure of cardiovascular determinants: a stable demographic foundation modulated by smoking exposure and complemented by non-linear lifestyle patterns involving diet, sleep, and physical activity. These results support the view that multifaceted behavioural combinations, rather than isolated factors, underpin the predictive landscape of lifestyle-based cardiovascular risk models.

#### SHAP Analysis

SHAP-based analyses provided a model-agnostic view of feature contributions to the predicted risk of outcome *(see Appendix table 11)*. Across all models, age and sex were consistently dominant, reaffirming their foundational role in long-term risk stratification. Smoking behaviours represented the most stable and influential modifiable domain, emphasising the cumulative impact of lifetime tobacco exposure and the robustness of smoking-related predictors across both linear and non-linear frameworks.

Tree-based models expanded upon these core predictors by identifying a broader range of lifestyle domains, including sleep quality, dietary patterns, and physical activity. Sleep behaviours appeared consistently across SHAP rankings, indicating strong and potentially non-linear associations that may be underrepresented in model-internal importance metrics. Dietary factors such as major dietary changes, bread type, cheese intake, and water consumption also contributed meaningfully, with SHAP systematically amplifying their influence relative to model-native scores. These patterns suggest that both sleep and diet convey clinically interpretable long-term risk signals even in the absence of biomarker data.

Physical activity appeared more in ensemble models, where SHAP distributed these effects more evenly, suggesting that model-native importance metrics may overweight activity features during tree splitting. Compared with internal measures, SHAP assigned greater influence to diet, sleep, and activity, revealing modifiable behavioural predictors that algorithm-specific metrics may underrepresent. Behavioural indicators such as mobile phone usage also contributed consistently across models, suggesting potential proxy roles for lifestyle routines or broader behavioural patterns. Collectively, SHAP analyses indicate that demographic and smoking exposures form the structural backbone of prediction, while broader lifestyle behaviours introduce clinically meaningful variability, offering a unified, model-agnostic perspective on behavioural determinants of cardiovascular risk.

#### LIME Analysis

LIME-based analyses provided complementary local explanations of model behaviour, highlighting both cross-model consistency and algorithm-specific variation (see Appendix table 12). Across all models, age and sex again dominated the rankings, re-affirming their stability as the foundational demographic determinants of cardiovascular risk. Beyond these fixed factors, smoking-related variables remained the most influential modifiable predictors, while sleep, dietary, and activity-related behaviours contributed secondary but interpretable signals.

LR explanations were concentrated on a small set of linear and intuitively interpretable lifestyle exposures, particularly direct and passive smoking, napping, and insomnia demonstrating that even simple additive models can recover clinically plausible behavioural patterns. In contrast, RF distributed its explanatory weight across a larger number of moderate contributors, including sleep and physical activity metrics as well as diverse behavioural indicators such as technology use, reflecting its tendency to partition data through many small, heterogeneous effects. XGBoost produced a broader and more balanced profile, integrating demographic and smoking factors with non-linear lifestyle interactions involving sleep quality, dietary habits, and activity frequency. This pattern illustrates XGBoost’s ability to combine core demographic drivers with nuanced lifestyle interactions, capturing subtle behavioural combinations that shape individual-level predictions beyond main effects.

Although the absolute magnitudes of local feature weights differed between event and non-event groups, their relative ordering remained stable across models, underscoring the robustness of the key determinants identified. Together, LIME results complement the global SHAP findings by clarifying how these determinants operate at the individual level: demographic and smoking factors form the structural basis of prediction, while broader lifestyle and behavioural domains modulate personalised risk through context-dependent interactions that differ across model classes.

Across all models, SHAP consistently highlighted age, sex, and smoking-related variables as the most influential predictors of cardiovascular risk, underscoring their robustness as core determinants. LIME, applied to the same feature subset used in SHAP analyses, confirmed these findings while redistributing relative weights in a sample-specific manner, giving greater prominence to context-dependent lifestyle behaviours at the individual level. LR relied on a focused set of additive behavioural exposures, RF distributed influence across many moderate contributors, and XGBoost combined demographic and smoking factors with sleep and dietary behaviours in ways that reflected complex non-linear interactions.

In Figure 3, we present an overall summary of feature importance across all models and interpretability methods. *Model Agreement* score, which reflects the consistency of feature rankings, highlights predictors that remain influential regardless of the modelling approach. A more detailed and fine-grained version of this aggregation is presented in *Appendix Table 13*. Together, these visual and tabular summaries show that demographic and smoking factors form the structural core of prediction, while broader lifestyle domains contribute model-specific and non-linear patterns that shape personalised risk.

**Figure 3.**
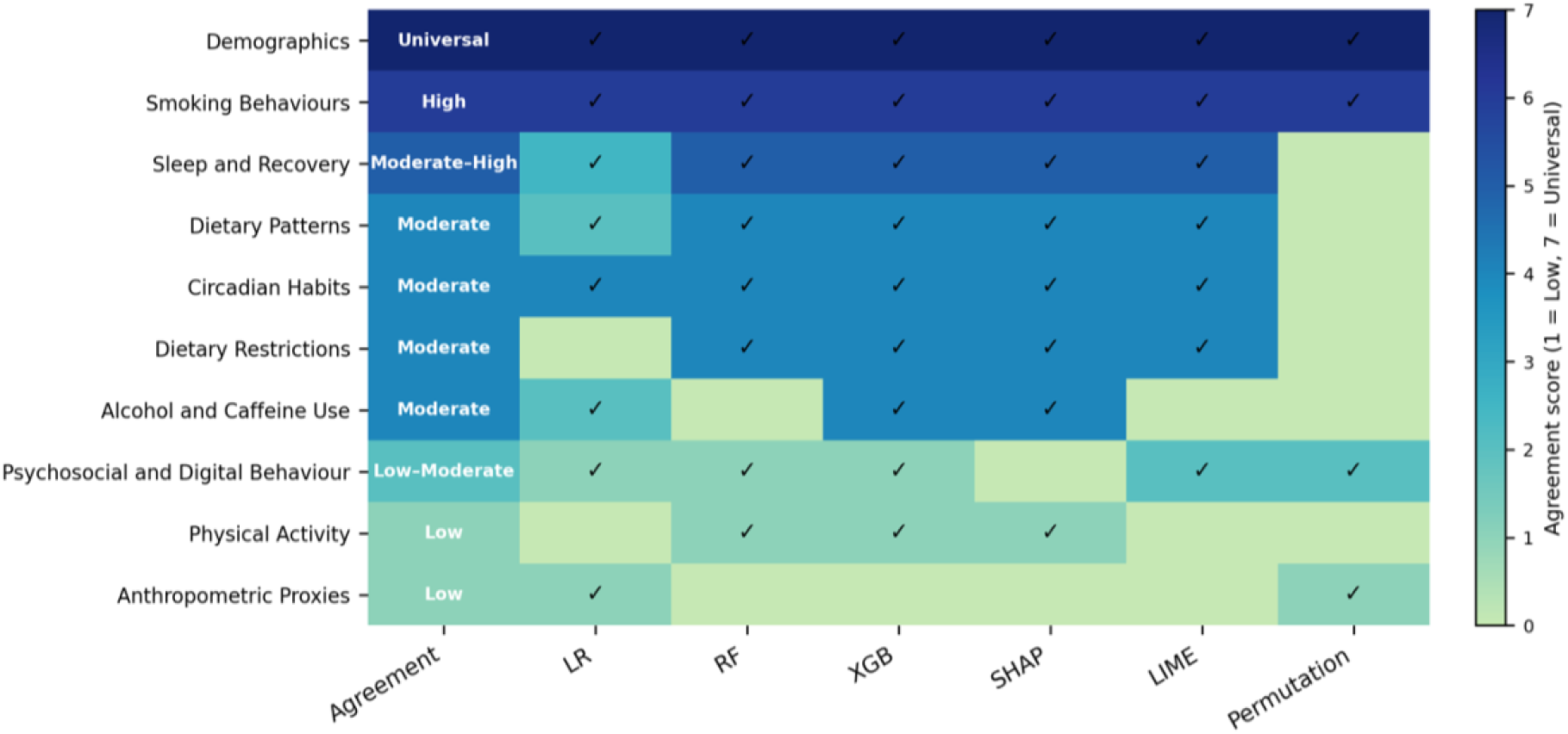
Summary of Lifestyle Feature Categories with Cross-model and Method Consistency

#### Subgroup Analysis

We selected four representative subgroups (youngest and oldest quartiles by sex: female Q1 to male Q4) spanning the full risk spectrum, from the lowest (female Q1, 2.4%) to the highest prevalence (male Q4, 20.4%). Figure 4 summarises discrimination across subgroups. Subgroups with ROC–AUC < 0.5 (e.g. XGBoost in female Q1) are not displayed for clarity but are reported in full in *Appendix Table 17*. Discrimination was broadly comparable (ROC-AUC 0.48–0.66; PR-AUC 0.02–0.25), but threshold-based metrics at the 0.5 cut-off revealed substantial disparities driven by prevalence. In low-risk groups, RF and XGBoost achieved high accuracy (>97%) but zero recall due to extreme class imbalance, whereas LR maintained low yet non-zero sensitivity, reflecting more stable calibration under sparse events. As prevalence increased, recall improved across models: RF reached ∼0.21 in high-risk males, and XGBoost provided a more balanced precision–recall trade-off. Calibration showed similar patterns: LR had the most stable Brier scores (0.026–0.202), RF tended to overestimate risk (0.164–0.231), and XGBoost remained intermediate. These findings indicate that subgroup prevalence has a major impact on recall and calibration, underscoring the need for subgroup-specific recalibration when deploying lifestyle-based models in heterogeneous populations Subgroup analyses were performed across all eight age–sex strata to evaluate demographic heterogeneity in model behaviour given adequate sample sizes within each stratum. Each subgroup was trained and evaluated separately using precomputed SHAP values to ensure comparability of feature attributions. Two representative groups were selected for detailed interpretation: young females (Sex = 0, Q1) and older males (Sex = 1, Q4) representing the lowest and highest cardiovascular risk profiles, respectively.

**Figure 4.**
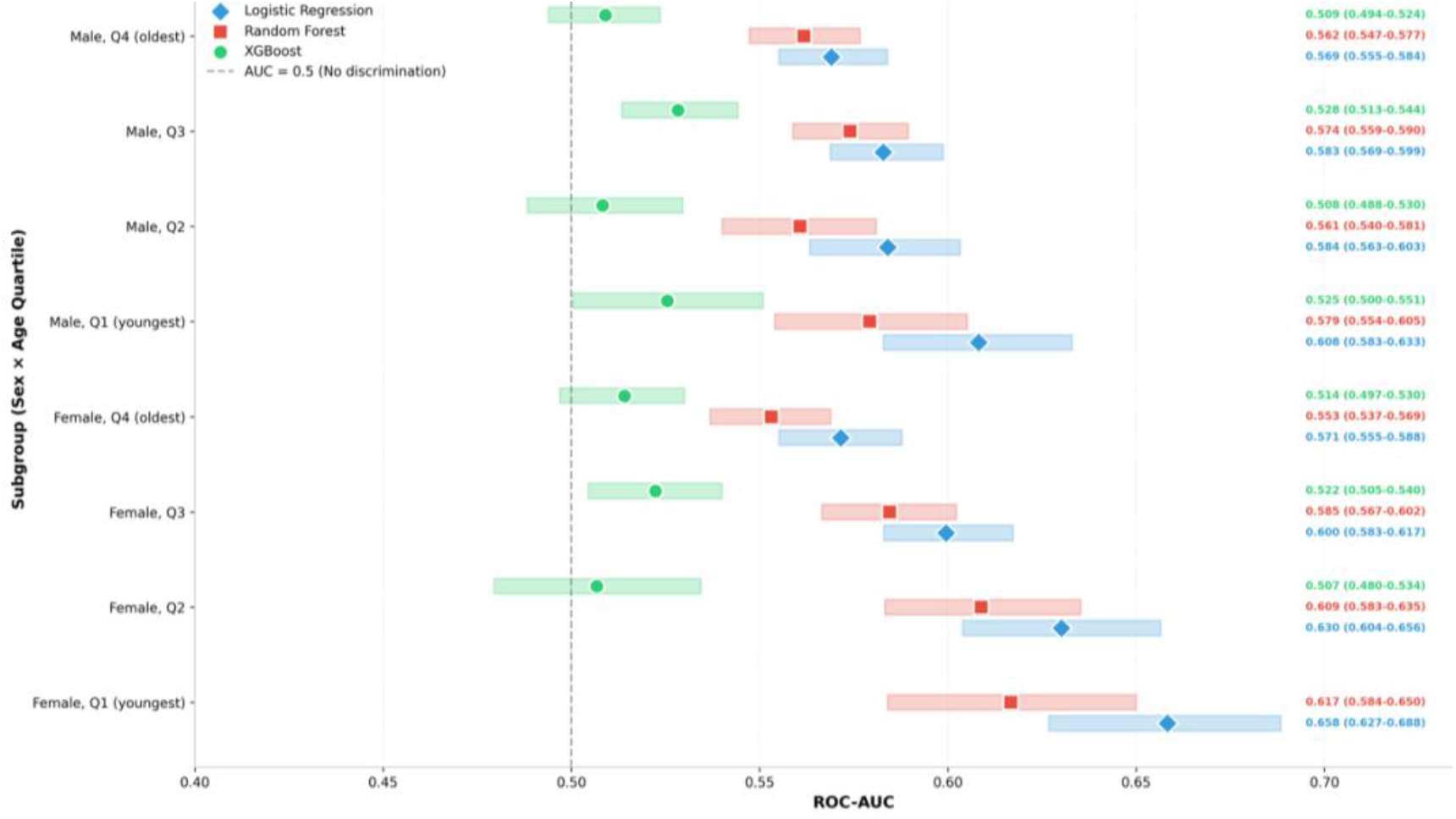
Subgroup-Specific Model Discrimination by Sex and Age Quartiles Results for subgroups where ROC-AUC < 0.5 were omitted from the plot for clarity; Age quartiles in cohort were defined as:Q1 = 37– 50 years, Q2 = 50–57 years, Q3 = 57–63 years, Q4 = 63–73 years.

Among younger females, model explanations were dominated by demographic and modifiable behavioural factors. LR emphasised smoking and sleep-related exposures, RF incorporated physical activity and daily routines, and XGBoost captured broader lifestyle domains including diet and social behaviour. These findings suggest that in early adulthood, proximal and modifiable behaviours are the primary drivers of cardiovascular risk. In contrast, older males exhibited feature importance dominated by cumulative exposures and entrenched habits. Across models, long-term smoking history, stable sleep patterns, and persistent dietary behaviours contributed most strongly, while short-term behavioural signals played a minor role.

When comparing all eight subgroups, age, sex, and smoking exposure consistently ranked as the leading predictors *(see Appendix section 12)*. However, the relative contribution of lifestyle domains shifted gradually with age from modifiable factors in younger groups to cumulative, long-term exposures in older adults highlighting an age-dependent transition in behavioural risk patterns relevant for targeted prevention.

## Discussion

### Principal Results

This study developed interpretable machine learning models to predict 10-year risks of CVD, HF, AF using lifestyle-based factors the UK Biobank. Integrating discrimination, calibration, and interpretability assessments, the models showed that behavioural data alone can meaningfully predict long-term cardiovascular risk. XGBoost achieved the highest discrimination (ROC-AUC = 0.752; PR-AUC = 0.230), followed closely by logistic regression (ROC-AUC = 0.748; PR-AUC = 0.225), while random forest showed weaker calibration. Logistic regression and XGBoost were therefore retained for interpretability analyses as they balanced accuracy and transparency.

Feature importance analyses identified both common and model-specific predictors. Age and sex were consistently dominant, while smoking, diet, and physical activity emerged as key modifiable factors. Smoking-related variables emphasised the need for early cessation support and behavioural screening, whereas dietary and activity factors highlighted opportunities for preventive counselling and lifestyle interventions. Subgroup analyses revealed performance heterogeneity: younger participants exhibited lower recall but better discrimination, while older groups showed the opposite trend. SHAP results further indicated that lifestyle and proximal behaviours were more influential in younger females, whereas cumulative exposures and parental longevity dominated in older males, reflecting an age-dependent shift in behavioural risk patterns.

Overall, interpretable ML uncovered actionable lifestyle determinants and subgroup-specific variations, bridging algorithmic prediction with behavioural prevention and supporting the development of transparent, data-driven risk assessment tools.

### Limitations

Key strengths include the use of a large, deeply phenotyped cohort (N = 459,142), robust cross-validation, and complementary interpretability analyses, which together enabled transparent evaluation of model performance and subgroup heterogeneity. But several limitations should also be acknowledged. First, the exclusion of clinical, biochemical, and genetic variables was intentional to ensure the model can be deployed in the general population without requiring laboratory tests or biomarker collection. This design improves accessibility and practical usability, but naturally limits predictive performance in high-risk or biomarker-rich clinical settings compared with models that incorporate clinical markers. Second, certain lifestyle variables are not routinely recorded across all clinical or digital systems, which may affect model transferability and real-world applicability. Third, external validation was not conduceted; generalisability was instead assessed through internal testing and subgroup-level interpretability. Future studies should prioritise temporal and cross-site validation across diverse healthcare systems to confirm robustness. Fourth, interpretability tools such as SHAP and LIME depend on local approximations and may yield unstable explanations when predictors are correlated, highlighting the need for methodological refinement and clinical validation. Fifth, the proposed models rely on a large set of lifestyle features, which may complicate direct clinical deployment; a smaller model using a limited set of routinely available variables may be needed to approximate the full model for real-world use. Finally, UK Biobank participants are typically healthier and less socio-demographically diverse than the wider UK population^27^, which may limit representativeness; however, the direction and relative importance of predictors remained consistent with findings from broader population studies^28^.

### Comparison with previous studies

Findings align with established evidence that age^29^, sex^30^, and smoking^31^ remain key determinants of CVD. Despite excluding clinical biomarkers, the models recovered classical risk patterns similar to those in Framingham, QRISK3, and PCE demonstrating that lifestyle data alone capture substantial exposure information. These results support the value of lifestyle-based frameworks in digital and community settings where laboratory measures are unavailable.

Feature-level effects were consistent with UK Biobank and meta-analytic findings: physical activity and certain dietary habits were protective, while smoking, alcohol, and processed meat increased risk. SHAP and LIME provided complementary interpretability at both global and local levels, linking model-derived explanations to epidemiological evidence, and enhancing clinical transparency.

## Conclusions

Lifestyle-based interpretable models offer a scalable and low-cost approach to cardiovascular risk assessment by leveraging key modifiable behaviours such as smoking patterns, alcohol intake, dietary habits, physical activity levels, and sleep characteristics. By highlighting these factors within model outputs, the approach supports earlier identification of at-risk individuals, enhances personalised risk communication, and facilitates behaviour-focused prevention through targeted counselling and intervention planning. In clinical contexts, they can complement established risk calculators by highlighting modifiable behaviours during consultations, especially for younger adults at low 10-year but high lifetime risk. In digital health applications, such models can deliver personalised feedback through mobile platforms or patient portals, supporting self-management and behavioural change. Future research should prioritise external validation across multi-regional and socioeconomically diverse cohorts, develop user-facing interfaces that translate model explanations into clinical language, and evaluate whether interpretability improves patient understanding, engagement, and adherence. This study also compared interpretability techniques across multiple models, demonstrating that SHAP, LIME, and model-specific importance measures converge on core lifestyle drivers while revealing model-dependent variation. Incorporating such interpretability assessments, including their performance across additional subgroups, will be important for promoting fairness and addressing health inequalities. Collectively, these directions move beyond algorithmic optimisation toward real-world, interpretable, and equitable cardiovascular prevention frameworks.

## Data Availability

The data used in this study are available from the UK Biobank under an approved application, but restrictions apply to the availability of these data, which were used under license for the current study and are not publicly available. Data are however available from the authors upon reasonable request and with permission of the UK Biobank.

https://www.ukbiobank.ac.uk/

## Acknowledgements

K.D. conceived the study idea and provided access to the original UK Biobank dataset. Y.F. curated and adapted the dataset for this analysis, performing variable selection and data preparation tailored to the study objectives. Y.F. designed and implemented all methodological procedures, conducted machine learning modelling and interpretability analyses, and drafted the manuscript. K.D. and H.K. reviewed the analytical approach, validated the interpretation, and provided critical feedback on the manuscript. All authors contributed to the refinement of the final version and approved the submitted manuscript. Y.F. is responsible for the integrity of the work as a whole.

Yuan Feng — Writing (original draft, editing);

Holger Kunz — Writing (review & editing);

Katarzyna Dziopa — Supervision, Validation, Writing (review & editing).

## Conflicts of Interest

none declared

## Abbreviations

CVD: Cardiovascular Diseases
HF: Heart Failure
AF: Atrial Fibrillation
AI: Artificial Intelligence
ML: Machine Learning
HDL: High-Density Lipoprotein
LDL: Low-Density Lipoprotein
BMI: Body Mass Index
SBP: Systolic Blood Pressure
DBP: Diastolic Blood Pressure
CRP: C-Reactive Protein
HbA1c: Glycated Hemoglobin
IQR: Interquartile Range
SD: Standard Deviation
mmHg: Millimetres of Mercury
mmol/L: Millimoles per Litre
µmol/L: Micromoles per Litre
SHAP: SHapley Additive exPlanations
LIME: Local Interpretable Model-agnostic Explanations
NHANES: National Health and Nutrition Examination Survey

## Multimedia Appendix

Appendix file

## Notes

### Competing Interest Statement

The authors have declared no competing interest.

### Funding Statement

This study did not receive any funding

### Author Declarations

The North West Multi-centre Research Ethics Committee of the United Kingdom gave ethical approval for the UK Biobank study. This research was conducted using the UK Biobank Resource under application number 12113.

