## Supplementary Appendix for "Interpretable Lifestyle-Based Machine Learning Models for Ten-Year Cardiovascular Risk Prediction using data from the UK Biobank"

#### Appendix Contents

**1 Final Feature ID Table (Field ID, Name, Type) 2**

**2 Features with Structural Missingness 4**

**3 Excluded Features (Not Matched) 5**

**4 Flowchart for Feature Selection 6**

**5 Model Performance and Tuning 7**

**7 Model Internal Top20 Feature Ranking 14**

**8 SHAP Top20 Feature Ranking 15**

**9 LIME Top20 Feature Ranking 16**

**10 Aggressive Summary of Cross-model and Cross-method Feature Categories 17**

**11 LIME Ranking for Features 18**

### 1 Final Feature ID Table (Field ID, Name, Type)

| ID | Field Name | Data Type |
|---|---|---|
| 1160 | Sleep duration | Integer |
| 1170 | Getting up in morning | Categorical (single) |
| 1180 | Morning/evening person (chronotype) | Categorical (single) |
| 1190 | Nap during day | Categorical (single) |
| 1200 | Sleeplessness / insomnia | Categorical (single) |
| 1210 | Snoring | Categorical (single) |
| 1220 | Daytime dozing / sleeping (narcolepsy) | Categorical (single) |
| 20160 | Ever smoked | Categorical (single) |
| 20162 | Pack years adult smoking as proportion of life span exposed to smoking | Continuous |
| 20161 | Pack years of smoking | Continuous |
| 20116 | Smoking status | Categorical (single) |
| 1239 | Current tobacco smoking | Categorical (single) |
| 1249 | Past tobacco smoking | Categorical (single) |
| 2644 | Light smokers, at least 100 smokes in lifetime | Categorical (single) |
| 3436 | Age started smoking in current smokers | Integer |
| 3446 | Type of tobacco currently smoked | Categorical (single) |
| 5959 | Previously smoked cigarettes on most/all days | Categorical (single) |
| 3456 | Number of cigarettes currently smoked daily | Integer |
| 6194 | Age stopped smoking cigarettes (current cigar/pipe or previous cigarette smoker) | Integer |
| 6183 | Number of cigarettes previously smoked daily (current cigar/pipe smokers) | Integer |
| 3466 | Time from waking to first cigarette | Categorical (single) |
| 3476 | Difficulty not smoking for 1 day | Categorical (single) |
| 3486 | Ever tried to stop smoking | Categorical (single) |
| 3496 | Wants to stop smoking | Categorical (single) |
| 3506 | Smoking compared to 10 years previous | Categorical (single) |
| 6158 | Why reduced smoking | Categorical (multiple) |
| 2867 | Age started smoking in former smokers | Integer |
| 2877 | Type of tobacco previously smoked | Categorical (single) |
| 2887 | Number of cigarettes previously smoked daily | Integer |
| 2897 | Age stopped smoking | Integer |
| 2907 | Ever stopped smoking for 6+ months | Categorical (single) |
| 6157 | Why stopped smoking | Categorical (multiple) |
| 2926 | Number of unsuccessful stop-smoking attempts | Integer |
| 2936 | Likelihood of resuming smoking | Categorical (single) |
| 1259 | Smoking/smokers in household | Categorical (single) |
| 1269 | Exposure to tobacco smoke at home | Integer |
| 1279 | Exposure to tobacco smoke outside home | Integer |
| 1289 | Cooked vegetable intake | Integer |
| 1299 | Salad / raw vegetable intake | Integer |
| 1309 | Fresh fruit intake | Integer |
| 1319 | Dried fruit intake | Integer |
| 1329 | Oily fish intake | Categorical (single) |
| 1339 | Non-oily fish intake | Categorical (single) |
| 1349 | Processed meat intake | Categorical (single) |
| 1359 | Poultry intake | Categorical (single) |

Continued on next page

| ID | Field Name | Data Type |
|---|---|---|
| 1369 | Beef intake | Categorical (single) |
| 1379 | Lamb/mutton intake | Categorical (single) |
| 1389 | Pork intake | Categorical (single) |
| 6144 | Never eat eggs, dairy, wheat, sugar | Categorical (multiple) |
| 1408 | Cheese intake | Categorical (single) |
| 1418 | Milk type used | Categorical (single) |
| 1428 | Spread type | Categorical (single) |
| 2654 | Non-butter spread type details | Categorical (single) |
| 1438 | Bread intake | Integer |
| 1448 | Bread type | Categorical (single) |
| 1458 | Cereal intake | Integer |
| 1468 | Cereal type | Categorical (single) |
| 1478 | Salt added to food | Categorical (single) |
| 1488 | Tea intake | Integer |
| 1498 | Coffee intake | Integer |
| 1508 | Coffee type | Categorical (single) |
| 1518 | Hot drink temperature | Categorical (single) |
| 1528 | Water intake | Integer |
| 1538 | Major dietary changes in the last 5 years | Categorical (single) |
| 1548 | Variation in diet | Categorical (single) |
| 22035 | Above moderate/vigorous recommendation | Categorical (single) |
| 22036 | Above moderate/vigorous/walking recommendation | Categorical (single) |
| 22032 | IPAQ activity group | Categorical (single) |
| 22038 | MET minutes per week for moderate activity | Continuous |
| 22039 | MET minutes per week for vigorous activity | Continuous |
| 22037 | MET minutes per week for walking | Continuous |
| 22040 | Summed MET minutes per week for all activity | Continuous |
| 22033 | Summed days activity | Integer |
| 22034 | Summed minutes activity | Integer |
| 1110 | Length of mobile phone use | Categorical (single) |
| 1120 | Weekly usage of mobile phone in last 3 months | Categorical (single) |
| 1130 | Hands-free device/speakerphone use | Categorical (single) |
| 1140 | Difference in mobile phone use vs. 2 years ago | Categorical (single) |
| 1150 | Usual side of head for mobile phone use | Categorical (single) |
| 2237 | Plays computer games | Categorical (single) |

#### 2 Features with Structural Missingness

| No. | Missing Rate | Field Name |
|---|---|---|
| 1 | 92.35% | Age started smoking in current smokers |
| 2 | 92.36% | Type of tobacco currently smoked |
| 3 | 99.5% | Previously smoked cigarettes on most/all days |
| 4 | 0.52% | Number of cigarettes currently smoked daily |
| 5 | 99.66% | Age stopped smoking cigarettes |
| 6 | 0.17% | Number of cigarettes previously smoked daily |
| 7 | 92.96% | Time from waking to first cigarette |
| 8 | 0.52% | Difficulty not smoking for 1 day |
| 9 | 92.39% | Ever tried to stop smoking |
| 10 | 92.36% | Wants to stop smoking |
| 11 | 92.37% | Smoking compared to 10 years previous |
| 12 | 76.39% | Age started smoking in former smokers |
| 13 | 76.39% | Type of tobacco previously smoked |
| 14 | 76.38% | Age stopped smoking |
| 15 | 76.39% | Ever stopped smoking for 6+ months |
| 16 | 76.38% | Number of unsuccessful stop-smoking attempts |
| 17 | 76.38% | Likelihood of resuming smoking |
| 18 | 7.65% | Smoking/smokers in household |

### 3 Excluded Features (Not Matched)

Some lifestyle-related features identified from the UK Biobank metadata were excluded due to one of two reasons: (1) the field was designated as a pilot variable and not available for general access; or (2) the corresponding field ID was not present in the current dataset extract, likely due to version differences or missing collection at the time of export. These ”not matched” features refer specifically to those present in the metadata but not found in the current working dataset after linkage by field ID.

Table 2: List of lifestyle-related features excluded due to not being matched in the working dataset.

| No. | Field Name | Category Path | Data Type |
|---|---|---|---|
| 1 | Light smokers, at least 100 smokes in lifetime (pilot) | Smoking – Touchscreen – Assessment Centre | Categorical (single) |
| 2 | Ever stopped smoking for 6+ months (pilot) | Smoking – Touchscreen – Assessment Centre | Categorical (single) |
| 3 | Why stopped smoking (pilot) | Smoking – Touchscreen – Assessment Centre | Categorical (single) |
| 4 | Age when last ate meat | Diet – Touchscreen – Assessment Centre | Integer |
| 5 | Never eat eggs, dairy, wheat, sugar (pilot) | Diet – Touchscreen – Assessment Centre | Categorical (multiple) |
| 6 | Spread type (pilot) | Diet – Touchscreen – Assessment Centre | Categorical (single) |
| 7 | Bread type/intake (pilot) | Diet – Touchscreen – Assessment Centre | Categorical (single) |
| 8 | Variation in diet (pilot) | Diet – Touchscreen – Assessment Centre | Categorical (single) |
| 9 | Time using mobile phone in last 3 months (pilot) | Electronic device use – Touchscreen – Assessment Centre | Categorical (single) |
| 10 | Regular hands-free device use (pilot) | Electronic device use – Touchscreen – Assessment Centre | Categorical (single) |
| 11 | Difference in mobile use vs. one year ago (pilot) | Electronic device use – Touchscreen – Assessment Centre | Categorical (single) |
| 12 | Internet user (pilot) | Electronic device use – Touchscreen – Assessment Centre | Categorical (single) |
| 13 | Willing to be contacted by email (pilot) | Electronic device use – Touchscreen – Assessment Centre | Categorical (single) |

#### 4 Flowchart for Feature Selection

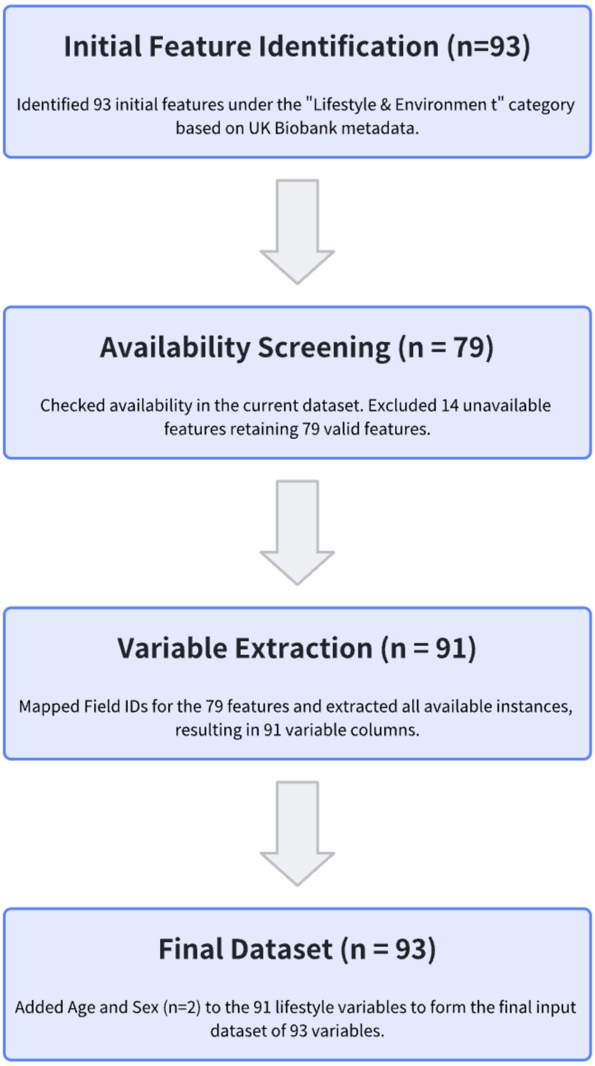

Figure 1: Flowchart of lifestyle-related feature selection from the UK Biobank dataset.

*Notes:* The feature selection process is illustrated in Appendix Figure 1. A total of 93 lifestyle and environment features were initially identified from the UK Biobank category-level metadata. 14 features were unavailable in the current data extract, leaving 79 valid features. After mapping the Field IDs and extracting all corresponding instances, 91 feature columns were obtained. Finally, age and sex were added as covariates, resulting in a final input dataset of 93 variables for analysis.

### 5 Model Performance and Tuning

#### 5.1 Logistic Regression

Table 3: Non-SMOTE Logistic Regression — 5-fold CV hyperparameter tuning

| Rank | C | Mean CV AUC (95% CI) |
|---|---|---|
| **1** | **0.01** | **0.7238 (0.722–0.725)** |
| 2 | 0.10 | 0.7237 (0.722–0.725) |
| 3 | 0.50 | 0.7237 (0.722–0.725) |
| 4 | 1.00 | 0.7237 (0.722–0.725) |
| 5 | 2.00 | 0.7237 (0.722–0.725) |

**Best model (C=0.01)** 5-fold CV ROC-AUC = 0.7238 (95% CI: 0.722–0.725).

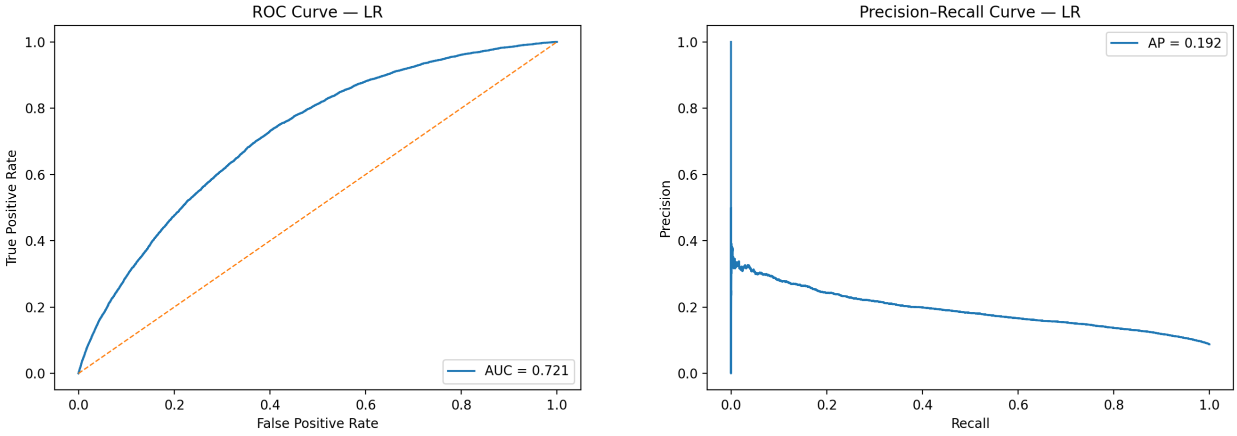

(a) Logistic Regression ROC

(b) Logistic Regression PR curve

Figure 2: LR performance curves.

Table 4: Threshold-dependent performance metrics for LR

| Threshold | Accuracy | Precision | Recall | F1 | Specificity | Balanced Acc. | Youden's J | TN | FP | FN | TP |
|---|---|---|---|---|---|---|---|---|---|---|---|
| 0.20 | 0.215 | 0.099 | 0.975 | 0.179 | 0.142 | 0.558 | 0.117 | 11884 | 71875 | 204 | 7866 |
| 0.25 | 0.285 | 0.106 | 0.955 | 0.190 | 0.221 | 0.588 | 0.175 | 18483 | 65276 | 365 | 7705 |
| 0.30 | 0.357 | 0.113 | 0.924 | 0.202 | 0.303 | 0.613 | 0.227 | 25367 | 58392 | 613 | 7457 |
| 0.35 | 0.431 | 0.122 | 0.887 | 0.215 | 0.387 | 0.637 | 0.274 | 32406 | 51353 | 910 | 7160 |
| 0.40 | 0.503 | 0.132 | 0.835 | 0.228 | 0.471 | 0.653 | 0.306 | 39480 | 44279 | 1332 | 6738 |
| 0.45 | 0.576 | 0.143 | 0.767 | 0.241 | 0.557 | 0.662 | 0.324 | 46692 | 37067 | 1882 | 6188 |
| 0.50 | 0.646 | 0.156 | 0.686 | 0.254 | 0.642 | 0.664 | 0.327 | 53748 | 30011 | 2538 | 5532 |
| 0.55 | 0.710 | 0.169 | 0.585 | 0.262 | 0.722 | 0.654 | 0.307 | 60480 | 23279 | 3346 | 4724 |

Table 5: Balanced Random Forest hyperparameter tuning

| Rank | Hyperparameters | Mean CV AUC (95% CI) |
|---|---|---|
| 1 | max_depth=None, leaf=50, split=100 | 0.718 (0.713–0.722) |
| 2 | max_depth=None, leaf=50, split=200 | 0.717 (0.712–0.721) |
| 3 | max_depth=None, leaf=100, split=100/200 | 0.715 (0.711–0.720) |
| 4 | max_depth=8, leaf=50, split=100 | 0.712 (0.708–0.717) |
| 5 | max_depth=8, leaf=50, split=200 | 0.712 (0.707–0.717) |
| 6 | max_depth=8, leaf=100, split=100/200 | 0.712 (0.707–0.717) |

*All models used 200 estimators, max_features=`sqrt`, and ccp_alpha=0.*

**Best model (Rank 1: max_depth=None, leaf=50, split=100)** achieved stable 5-fold CV performance: ROC-AUC = 0.718 (0.713–0.722).

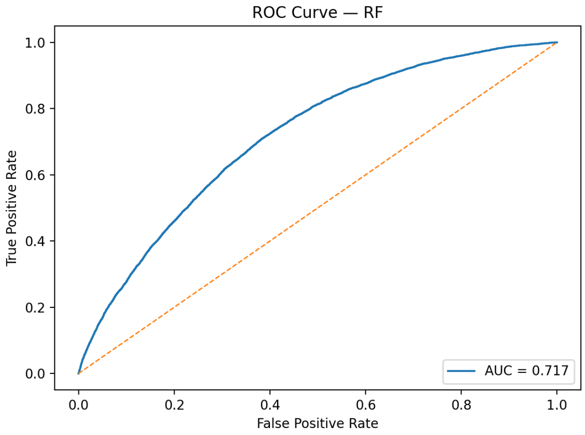

(a) Random Forest ROC

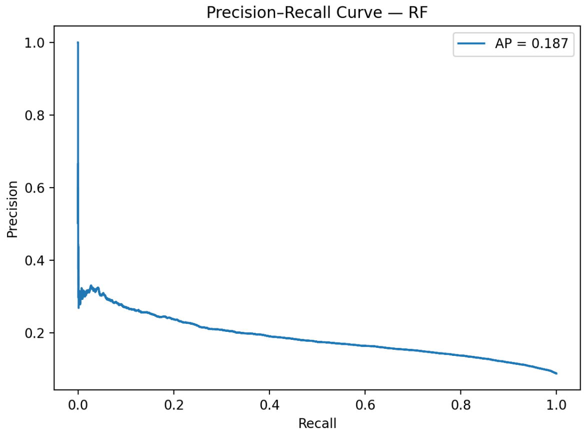

(b) Random Forest PR curve

Figure 3: RF performance curves.

#### 5.2 Random Forest

Table 6: Threshold-dependent performance metrics for **Random Forest**

| Threshold | Accuracy | Precision | Recall | F1 | Specificity | Balanced Acc. | Youden's J | TN | FP | FN | TP |
|---|---|---|---|---|---|---|---|---|---|---|---|
| 0.200 | 0.104 | 0.089 | 0.998 | 0.164 | 0.018 | 0.508 | 0.016 | 1481 | 82278 | 17 | 8053 |
| 0.250 | 0.156 | 0.094 | 0.991 | 0.171 | 0.076 | 0.533 | 0.066 | 6328 | 77431 | 74 | 7996 |
| 0.300 | 0.237 | 0.101 | 0.969 | 0.182 | 0.166 | 0.568 | 0.135 | 13921 | 69838 | 248 | 7822 |
| 0.350 | 0.333 | 0.111 | 0.936 | 0.198 | 0.275 | 0.606 | 0.212 | 23064 | 60695 | 513 | 7557 |
| 0.400 | 0.433 | 0.122 | 0.881 | 0.214 | 0.389 | 0.635 | 0.271 | 32624 | 51135 | 960 | 7110 |
| 0.450 | 0.535 | 0.137 | 0.808 | 0.234 | 0.508 | 0.658 | 0.316 | 42567 | 41192 | 1552 | 6518 |
| 0.500 | 0.635 | 0.153 | 0.693 | 0.250 | 0.630 | 0.661 | 0.322 | 52746 | 31013 | 2481 | 5589 |
| 0.550 | 0.728 | 0.171 | 0.543 | 0.260 | 0.746 | 0.644 | 0.289 | 62498 | 21261 | 3692 | 4378 |

Table 7: XGBoost (Non-SMOTE) — 5-fold CV hyperparameter tuning

| Rank | Hyperparameters | Mean CV AUC (95% CI) |
|---|---|---|
| 1 | lr=0.1, depth=3, trees=200 | 0.728 (0.726–0.730) |
| 2 | lr=0.05, depth=3, trees=200 | 0.727 (0.725–0.729) |
| 3 | lr=0.05, depth=6, trees=200 | 0.726 (0.725–0.728) |
| 4 | lr=0.1, depth=6, trees=200 | 0.721 (0.719–0.723) |

*All models used subsample=0.8, colsample_bytree=0.8.*

**Best model (Rank 1: lr=0.1, depth=3, trees=200)** achieved stable 5-fold CV performance: ROC-AUC = 0.728 (0.726–0.730).

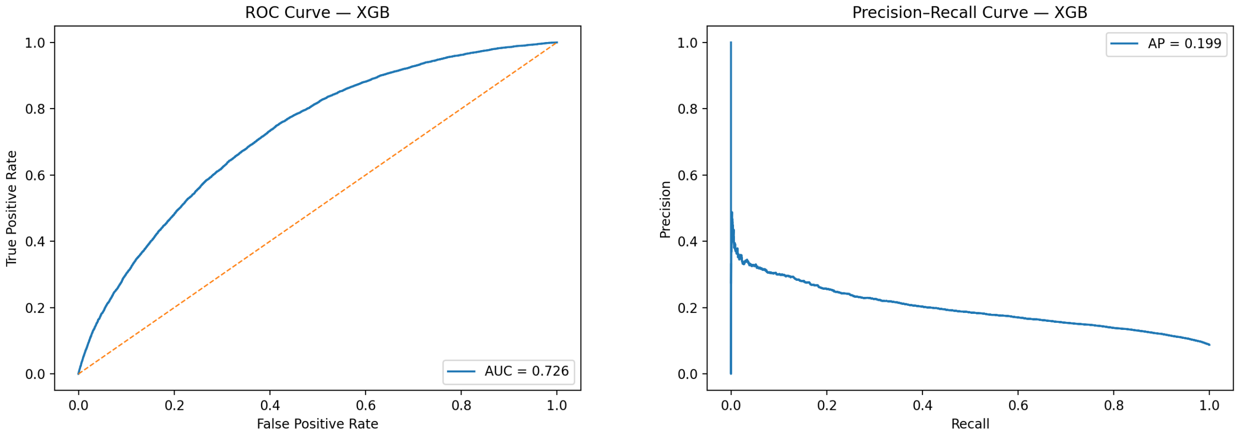

(a) XGBoost ROC

(b) XGBoost Precision–Recall curve

Figure 4: XGBoost performance curves.

#### 5.3 XGBoost

Table 8: Threshold-dependent performance metrics for **XGBoost**

| Threshold | Accuracy | Precision | Recall | F1 | Specificity | Balanced Acc. | Youden's J | TN | FP | FN | TP |
|---|---|---|---|---|---|---|---|---|---|---|---|
| 0.200 | 0.233 | 0.101 | 0.972 | 0.182 | 0.162 | 0.567 | 0.134 | 13549 | 70210 | 222 | 7848 |
| 0.250 | 0.303 | 0.108 | 0.950 | 0.193 | 0.241 | 0.596 | 0.191 | 20187 | 63572 | 400 | 7670 |
| 0.300 | 0.369 | 0.115 | 0.921 | 0.204 | 0.316 | 0.618 | 0.237 | 26437 | 57322 | 636 | 7434 |
| 0.350 | 0.437 | 0.123 | 0.885 | 0.216 | 0.394 | 0.639 | 0.279 | 32970 | 50789 | 928 | 7142 |
| 0.400 | 0.505 | 0.133 | 0.839 | 0.230 | 0.473 | 0.656 | 0.312 | 39640 | 44119 | 1299 | 6771 |
| 0.450 | 0.573 | 0.144 | 0.776 | 0.242 | 0.554 | 0.665 | 0.330 | 46385 | 37374 | 1804 | 6266 |
| 0.500 | 0.639 | 0.155 | 0.696 | 0.253 | 0.634 | 0.665 | 0.330 | 53103 | 30656 | 2454 | 5616 |
| 0.550 | 0.704 | 0.170 | 0.607 | 0.265 | 0.714 | 0.660 | 0.320 | 59767 | 23992 | 3173 | 4897 |

##### 5.4 Overall Summary Table

Table 9: Performance comparison of Logistic Regression, Random Forest, and XGBoost on the test set.

| Metric | Logistic Regression | Random Forest | XGBoost |
|---|---|---|---|
| **Cross-validation (5-fold)** | | | |
| CV ROC-AUC (mean ± sd) | 0.724 ± 0.0016 | 0.718 ± 0.0023 | 0.728 ± 0.0018 |
| Param combinations | 5 | 8 | 4 |
| **Test performance** | | | |
| ROC-AUC (95% CI) | [0.716–0.726] | [0.711–0.722] | [0.720–0.731] |
| PR-AUC | 0.192 | 0.187 | 0.199 |
| **Best-F1 threshold** | | | |
| Threshold | 0.55 | 0.55 | 0.55 |
| Accuracy | 0.710 | 0.728 | 0.704 |
| Precision | 0.169 | 0.171 | 0.170 |
| Recall | 0.585 | 0.543 | 0.607 |
| F1 | 0.262 | 0.260 | 0.265 |

### 6 SHAP Plot

#### 6.1 Logistic Regression

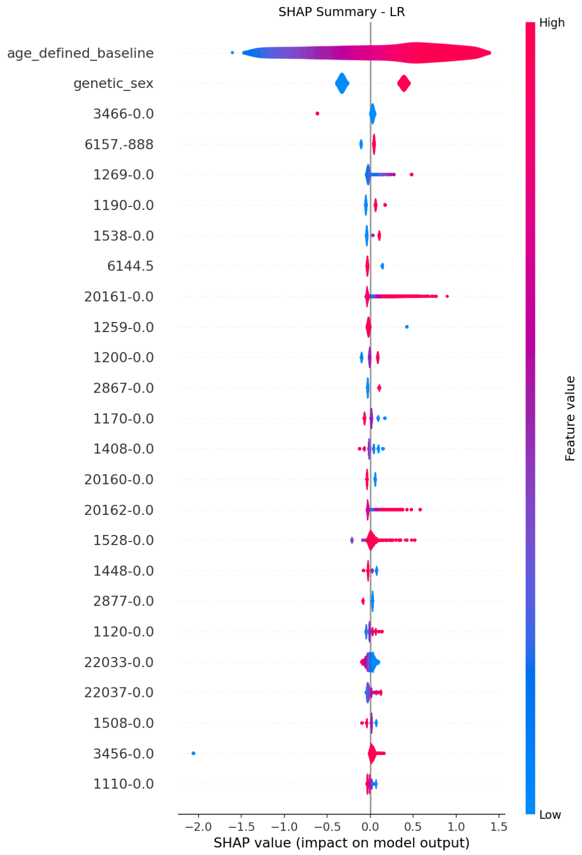

(a) SHAP Plot for Logistic Regression

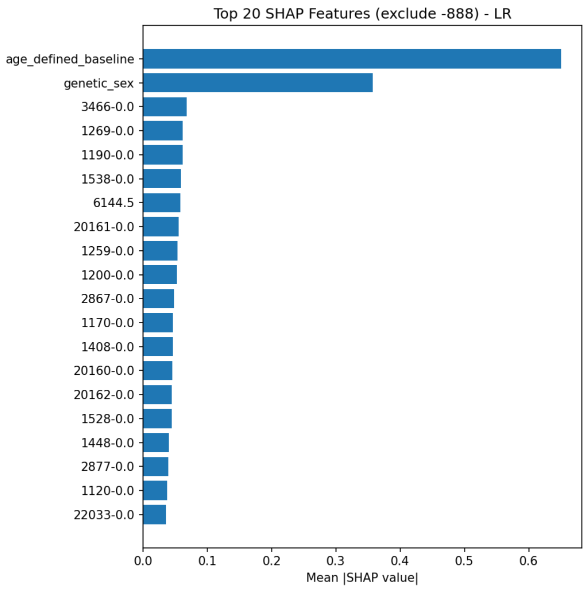

(b) Forest Plot for Logistic Regression

Figure 5: SHAP plot for Logistic Regression.

#### 6.2 Random Forest

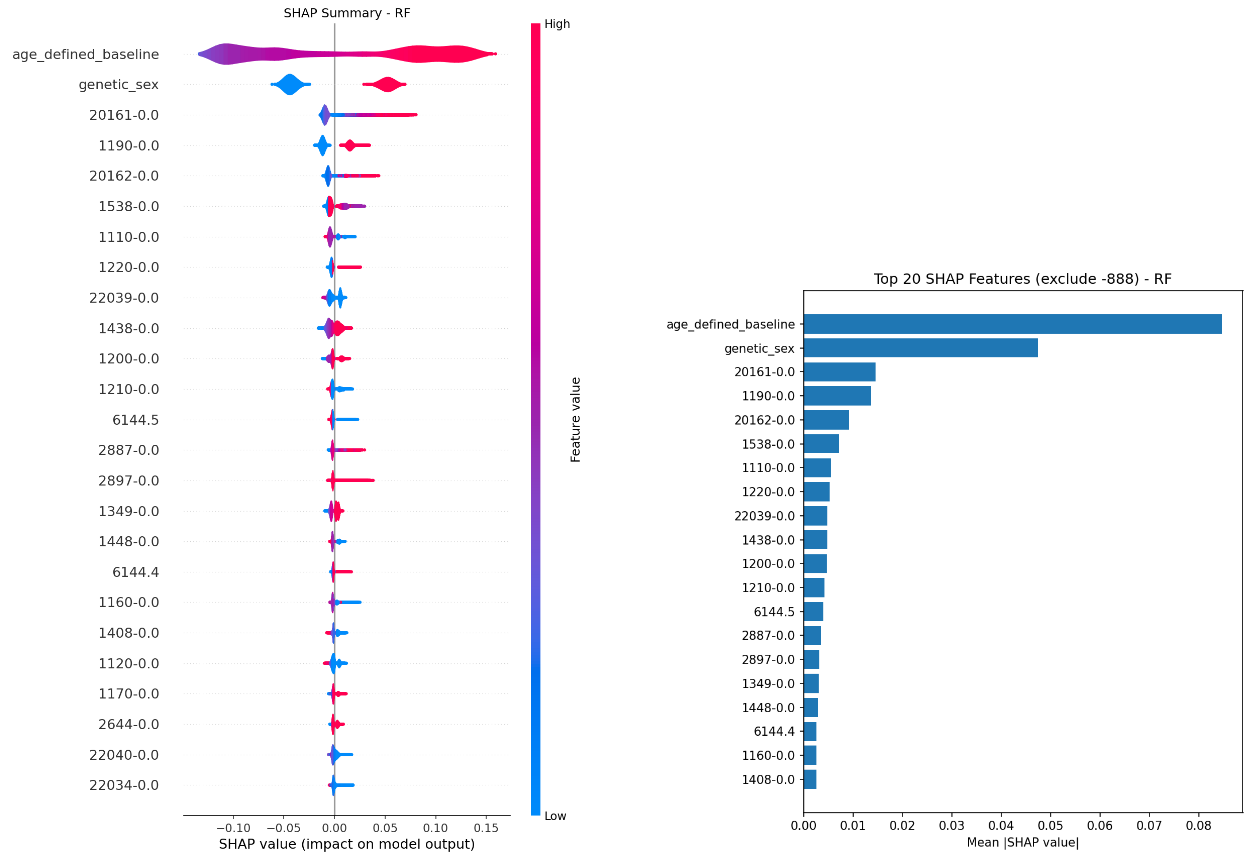

(a) SHAP Plot for Random Forest

(b) Forest Plot for Random Forest

Figure 6: SHAP plot for Random Forest.

#### 6.3 XGBoost

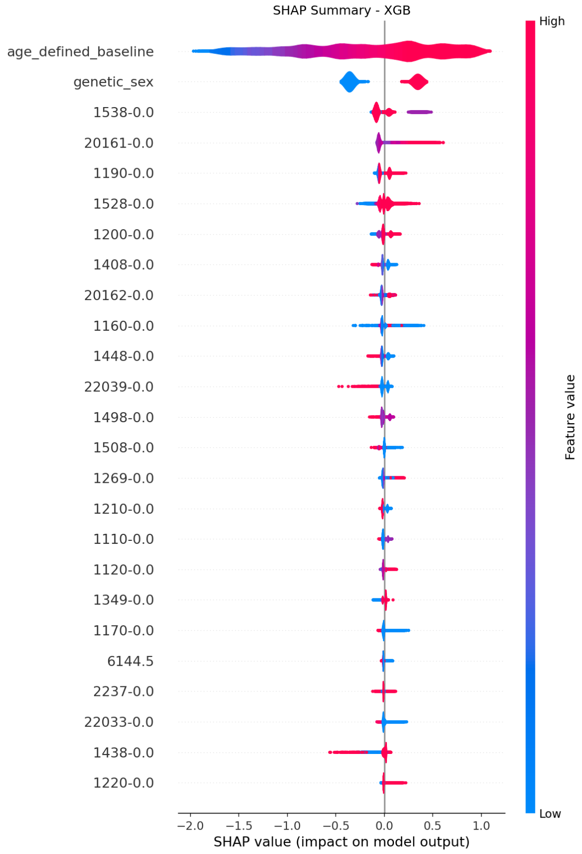

(a) SHAP Plot for XGBoost

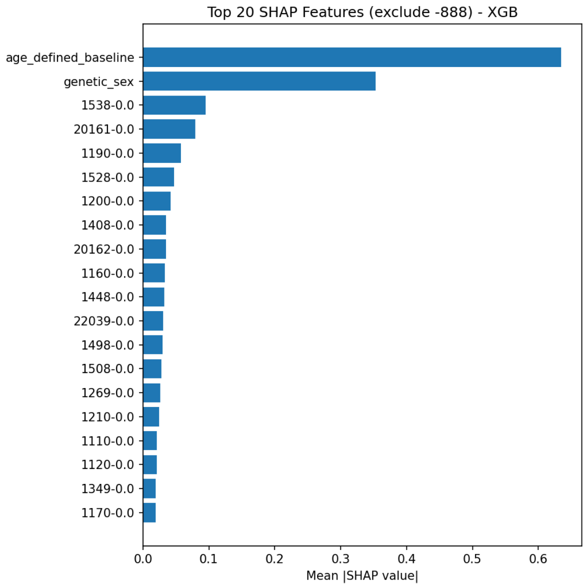

(b) Forest Plot for XGBoost

Figure 7: SHAP plot for XGBoost.

### 7 Model Internal Top20 Feature Ranking

Table 10: Top-20 substantive features ranked by importance across models.

| Rank | Logistic Regression (Permutation) | Random Forest (Gini) | XGBoost (Gain) |
|---|---|---|---|
| 1 | Age at baseline (age_defined_baseline) | Age at baseline (age_defined_baseline) | Age at baseline (age_defined_baseline) |
| 2 | Sex (genetic_sex) | Sex (genetic_sex) | Sex (genetic_sex) |
| 3 | Getting up in morning (1170) | Pack years of smoking (20161) | Major dietary changes in the last 5 years (1538) |
| 4 | Smoking/smokers in household (1259) | Nap during day (1190) | Pack years of smoking (20161) |
| 5 | Exposure to tobacco smoke at home (1269) | Pack years as proportion of lifespan (20162) | Nap during day (1190) |
| 6 | Weekly usage of mobile phone in last 3 months (1120) | Summed MET minutes per week for all activity (22040) | Water intake (1528) |
| 7 | Pack years of smoking (20161) | Bread intake (1438) | Sleeplessness / insomnia (1200) |
| 8 | Time from waking to first cigarette (3466) | Summed minutes of activity (22034) | Cheese intake (1408) |
| 9 | Water intake (1528) | Number of cigarettes previously smoked daily (2887) | Pack years adult smoking as proportion of life span exposed (20162) |
| 10 | Bread type (1448) | Vigorous MET minutes/week (22039) | Sleep duration (1160) |
| 11 | Pack years as proportion of lifespan (20162) | Age stopped smoking (2897) | Bread type (1448) |
| 12 | Never eat eggs, dairy, wheat, sugar (6144; multi-select) | Moderate MET minutes/week (22038) | MET minutes per week for vigorous activity (22039) |
| 13 | Wants to stop smoking (3496) | Length of mobile phone use (1110) | Coffee intake (1498) |
| 14 | Plays computer games (2237) | Walking MET minutes/week (22037) | Coffee type (1508) |
| 15 | Cheese intake (1408) | Summed days of activity (22033) | Exposure to tobacco smoke at home (1269) |
| 16 | Age started smoking in former smokers (2867) | Major dietary changes in the last 5 years (1538) | Snoring (1210) |
| 17 | Type of tobacco previously smoked (2877) | Past tobacco smoking (1249) | Length of mobile phone use (1110) |
| 18 | Coffee intake (1498) | Weekly mobile phone usage (1120) | Weekly usage of mobile phone in last 3 months (1120) |
| 19 | Age stopped smoking (2897) | Daytime dozing / narcolepsy (1220) | Processed meat intake (1349) |
| 20 | Current tobacco smoking (1239) | Processed meat intake (1349) | Getting up in morning (1170) |

*Note:* Feature IDs are mapped to descriptive UK Biobank variable names using the metadata table. This ranking excludes missing-indicator variables and artificial codes (e.g., "-888").

#### 8 SHAP Top20 Feature Ranking

Table 11: Top-20 substantive features ranked by SHAP importance across models (NO missing)

| Rank | Logistic Regression (SHAP) | Random Forest (SHAP) | XGBoost (SHAP) |
|---|---|---|---|
| 1 | Age at baseline (age_defined_baseline) | Age at baseline (age_defined_baseline) | Age at baseline (age_defined_baseline) |
| 2 | Sex (genetic_sex) | Sex (genetic_sex) | Sex (genetic_sex) |
| 3 | Time from waking to first cigarette (3466) | Pack years of smoking (20161) | Major dietary changes in the last 5 years (1538) |
| 4 | Exposure to tobacco smoke at home (1269) | Nap during day (1190) | Pack years of smoking (20161) |
| 5 | Nap during day (1190) | Pack years adult smoking as proportion of lifespan (20162) | Water intake (1528) |
| 6 | Major dietary changes in the last 5 years (1538) | Major dietary changes in the last 5 years (1538) | Sleeplessness / insomnia (1200) |
| 7 | Never eat eggs, dairy, wheat, sugar (multi-select) (6144.5) | Length of mobile phone use (1110) | Cheese intake (1408) |
| 8 | Pack years of smoking (20161) | Daytime dozing / sleeping (narcolepsy) (1220) | Bread type (1448) |
| 9 | Smoking/smokers in household (1259) | MET minutes/week for vigorous activity (22039) | MET minutes/week for vigorous activity (22039) |
| 10 | Sleeplessness / insomnia (1200) | Bread intake (1438) | Coffee intake (1498) |
| 11 | Age started smoking in former smokers (2867) | Sleeplessness / insomnia (1200) | Poultry intake (1508) |
| 12 | Getting up in morning (1170) | Snoring (1210) | Exposure to tobacco smoke at home (1269) |
| 13 | Cheese intake (1408) | Never eat eggs, dairy, wheat, sugar (multi-select) (6144.5) | Snoring (1210) |
| 14 | Ever smoked (20160) | Number of cigarettes previously smoked daily (2887) | Length of mobile phone use (1110) |
| 15 | Pack years adult smoking as proportion of lifespan (20162) | Age stopped smoking (2897) | Weekly usage of mobile phone in last 3 months (1120) |
| 16 | Water intake (1528) | Processed meat intake (1349) | Processed meat intake (1349) |
| 17 | Bread type (1448) | Bread type (1448) | Getting up in morning (1170) |
| 18 | Type of tobacco previously smoked (2877) | Never eat eggs, dairy, wheat, sugar (multi-select) (6144.4) | Sleep duration (1160) |
| 19 | Weekly usage of mobile phone in last 3 months (1120) | Sleep duration (1160) | Cheese intake (1408) |
| 20 | Summed days activity (22033) | Cheese intake (1408) | Bread intake (1438) |

*Note:* Items are ordered by mean absolute SHAP value within each model. Descriptive names are mapped from your metadata; base IDs are retained in parentheses. Codes such as 6144.5/6144.4 denote option-level encodings under the multi-select field *Never eat eggs, dairy, wheat, sugar* (ID 6144).

### 9 LIME Top20 Feature Ranking

Table 12: Top-20 substantive features ranked by LIME across models.

| Rank | Logistic Regression (LIME) | Random Forest (LIME) | XGBoost (LIME) |
|---|---|---|---|
| 1 | Age at baseline (age_defined_baseline) | Age at baseline (age_defined_baseline) | Age at baseline (age_defined_baseline) |
| 2 | Sex (genetic_sex) | Sex (genetic_sex) | Sex (genetic_sex) |
| 3 | Exposure to tobacco smoke at home (1269) | Pack years of smoking (20161) | Major dietary changes in the last 5 years (1538) |
| 4 | Nap during day (1190) | Nap during day (1190) | Sleeplessness / insomnia (1200) |
| 5 | Pack years of smoking (20161) | Pack years adult smoking as proportion of life span exposed (20162) | Pack years of smoking (20161) |
| 6 | Sleeplessness / insomnia (1200) | Number of cigarettes previously smoked daily (2887) | Sleep duration (1160) |
| 7 | Bread type (1448) | Length of mobile phone use (1110) | Water intake (1528) |
| 8 | Getting up in morning (1170) | Daytime dozing / sleeping (narcolepsy) (1220) | Nap during day (1190) |
| 9 | Smoking/smokers in household (1259) | Bread intake (1438) | Coffee type (1508) |
| 10 | Water intake (1528) | Sleeplessness / insomnia (1200) | Getting up in morning (1170) |
| 11 | Pack years adult smoking as proportion of life span exposed (20162) | MET minutes per week for vigorous activity (22039) | Cheese intake (1408) |
| 12 | Major dietary changes in the last 5 years (1538) | Age stopped smoking (2897) | Pack years adult smoking as proportion of life span exposed (20162) |
| 13 | Cheese intake (1408) | Never eat eggs, dairy, wheat, sugar (pilot) (6144; subfield 6144.4) | Exposure to tobacco smoke at home (1269) |
| 14 | Ever smoked (20160) | Processed meat intake (1349) | MET minutes per week for vigorous activity (22039) |
| 15 | Weekly usage of mobile phone in last 3 months (1120) | Snoring (1210) | Snoring (1210) |
| 16 | Summed days activity (22033) | Sleep duration (1160) | Length of mobile phone use (1110) |
| 17 | Time from waking to first cigarette (3466) | Bread type (1448) | Bread type (1448) |
| 18 | Type of tobacco previously smoked (2877) | Cheese intake (1408) | Weekly usage of mobile phone in last 3 months (1120) |
| 19 | Age started smoking in former smokers (2867) | Major dietary changes in the last 5 years (1538) | Processed meat intake (1349) |
| 20 | Never eat eggs, dairy, wheat, sugar (pilot) (6144; subfield 6144.5) | Never eat eggs, dairy, wheat, sugar (pilot) (6144; subfield 6144.5) | Coffee intake (1498) |

*Notes:* 1) LIME importance is the mean absolute local weight aggregated across individuals. 2) XGBoost LIME results were provided at rule level and have been aggregated to the feature level. 3) Fields 6144.4 and 6144.5 are subfields under 6144

### 10 Aggressive Summary of Cross-model and Cross-method Feature Categories

Table 13: Cross-model and cross-method feature category consistency, summarising how individual UK Biobank variables map into broader lifestyle constructs.

| **Category** | **Example variables** | **Summary of how top-ranked features map to category** | **Agreement level** |
|---|---|---|---|
| Demographics | Age, Sex | Age and sex consistently rank at the very top across all internal model metrics and SHAP/LIME, indicating strong, stable and universally reproducible effects. | Universal |
| Smoking Behaviours | Pack years, Current smoking, Passive smoke exposure, Age started/stopped, Tobacco type | Smoking indicators repeatedly appear among the highest-ranked features for all models. Tree models emphasise cumulative exposure, while LR captures timing and quitting, together forming a coherent *lifetime tobacco burden.* | High |
| Sleep and Recovery | Insomnia, Sleep duration, Napping, Getting-up time, Daytime dozing, Snoring | Sleep-related behaviours rank strongly in RF/XGB and more weakly in LR, suggesting mainly non-linear effects better captured by tree-based models. | Moderate–High |
| Dietary Patterns | Bread type, Cheese, Processed meat, Water intake, Dietary changes, Coffee type/intake, Poultry | Core dietary variables, especially major dietary change and fat/salt-rich foods, appear consistently in RF/XGB and SHAP/LIME; LR shows similar but attenuated patterns. | Moderate |
| Circadian Habits | Getting-up time, Napping frequency, Daytime dozing | Circadian timing and daytime sleepiness features repeatedly occur as SHAP/LIME rules with clear thresholds across models, indicating stable but modest contributions. | Moderate |
| Dietary Restrictions | Never eat eggs, dairy, wheat, sugar | Strict avoidance behaviours enter RF/XGB internal rankings and SHAP/LIME explanations, capturing a distinct "restrictive diet / health-consciousness" pattern. | Moderate |
| Psychosocial and Digital Behaviour | Mobile phone use, Computer gaming | Digital-usage variables appear across SHAP/LIME and RF/XGB, but rankings and direction vary by model, indicating context-dependent and less stable effects. | Low–Moderate |
| Alcohol and Caffeine Use | Coffee intake/type, Alcohol frequency/amount | Caffeine-related variables and alcohol frequency are highlighted most strongly by XGB and to a lesser extent by RF, with weaker and less consistent effects in LR. | Moderate |
| Physical Activity | Vigorous/moderate/walking activity (MET-min/week), Days active per week | Activity measures show clear non-linear dose–response effects in RF/XGB (internal importance and SHAP/LIME) but contribute little to LR permutation importance. | Low |
| Anthropometric Proxies | BMI, Self-rated health, Body weight perception | Anthropometric and general health measures have modest influence in LR but rarely enter RF/XGB top rankings, suggesting limited extra value once detailed lifestyle variables are included. | Low |

*Note.* Agreement levels and category mappings were derived through manual synthesis of all three importance tables (Table10, 11, 12). Each lifestyle variable was coded into categories and assessed for its recurrence, ranking stability, and cross-method consistency. The final levels represent qualitative frequency-based judgments of how reliably each category contributes across models and explanation methods.

### 11 LIME Ranking for Features

#### 11.1 Logistic Regression

Table 14: LIME aggregated explanations for **Logistic Regression** (*per-side* Top-20 by Mean |weight|), stratified by outcome ($y = 0$ vs. $y = 1$).

| | **Outcome = 0 (Non-event)** | | | | | | **Outcome = 1 (Event)** | | | | | |
|---|---|---|---|---|---|---|---|---|---|---|---|---|
| **Idx** | **Feature (thresholded)** | **Count** | **Mean** | **Std** | $\lvert w \rvert$ | **P(+)** | **Feature (thresholded)** | **Count** | **Mean** | **Std** | $\lvert w \rvert$ | **P(+)** |
| 1 | age_defined_baseline $\leq$ 53.00 | 3126 | -0.271 | 0.013 | 0.271 | 0.000 | age_defined_baseline $\leq$ 53.00 | 1283 | -0.275 | 0.015 | 0.275 | 0.000 |
| 2 | age_defined_baseline > 65.00 | 1006 | 0.238 | 0.006 | 0.238 | 1.000 | age_defined_baseline > 65.00 | 2111 | 0.239 | 0.007 | 0.239 | 1.000 |
| 3 | 0.00 < genetic_sex $\leq$ 1.00 | 3426 | 0.150 | 0.006 | 0.150 | 1.000 | 0.00 < genetic_sex $\leq$ 1.00 | 4771 | 0.150 | 0.007 | 0.150 | 1.000 |
| 4 | genetic_sex $\leq$ 0.00 | 4574 | -0.149 | 0.005 | 0.149 | 0.000 | genetic_sex $\leq$ 0.00 | 3229 | -0.150 | 0.005 | 0.150 | 0.000 |
| 5 | 60.00 < age_defined_baseline $\leq$ 65.00 | 1786 | 0.123 | 0.008 | 0.123 | 1.000 | 60.00 < age_defined_baseline $\leq$ 65.00 | 2628 | 0.124 | 0.009 | 0.124 | 1.000 |
| 6 | 1269-0.0 > 0.00 | 960 | 0.059 | 0.006 | 0.059 | 1.000 | 1269-0.0 $\leq$ 0.00 | 6667 | -0.059 | 0.006 | 0.059 | 0.000 |
| 7 | 1269-0.0 $\leq$ 0.00 | 7040 | -0.058 | 0.005 | 0.058 | 0.000 | 1269-0.0 > 0.00 | 1333 | 0.059 | 0.006 | 0.059 | 1.000 |
| 8 | 1190-0.0 > 2.00 | 368 | 0.050 | 0.008 | 0.050 | 1.000 | 1190-0.0 > 2.00 | 589 | 0.050 | 0.008 | 0.050 | 1.000 |
| 9 | 1190-0.0 $\leq$ 1.00 | 4665 | -0.049 | 0.004 | 0.049 | 0.000 | 1190-0.0 $\leq$ 1.00 | 3789 | -0.049 | 0.004 | 0.049 | 0.000 |
| 10 | 20161-0.0 > 12.00 | 1520 | 0.044 | 0.005 | 0.044 | 1.000 | 20161-0.0 > 12.00 | 2455 | 0.044 | 0.005 | 0.044 | 1.000 |
| 11 | 53.00 < age_defined_baseline $\leq$ 60.00 | 2082 | -0.041 | 0.004 | 0.041 | 0.000 | 53.00 < age_defined_baseline $\leq$ 60.00 | 1978 | -0.040 | 0.005 | 0.040 | 0.000 |
| 12 | 1.00 < 1190-0.0 $\leq$ 2.00 | 2967 | 0.039 | 0.004 | 0.039 | 1.000 | 1.00 < 1190-0.0 $\leq$ 2.00 | 3622 | 0.040 | 0.004 | 0.040 | 1.000 |
| 13 | 20161-0.0 $\leq$ 0.00 | 5597 | -0.033 | 0.004 | 0.033 | 0.000 | 20161-0.0 $\leq$ 0.00 | 4569 | -0.033 | 0.004 | 0.033 | 0.000 |
| 14 | 20162-0.0 > 0.29 | 1578 | 0.031 | 0.004 | 0.031 | 1.000 | 20162-0.0 > 0.29 | 2418 | 0.031 | 0.004 | 0.031 | 1.000 |
| 15 | 2.00 < 1200-0.0 $\leq$ 3.00 | 2184 | 0.028 | 0.004 | 0.028 | 1.000 | 2.00 < 1200-0.0 $\leq$ 3.00 | 2674 | 0.028 | 0.004 | 0.028 | 1.000 |
| 16 | 1448-0.0 $\leq$ 1.00 | 2054 | 0.028 | 0.004 | 0.028 | 1.000 | 1448-0.0 $\leq$ 1.00 | 2392 | 0.028 | 0.004 | 0.028 | 1.000 |
| 17 | 1200-0.0 $\leq$ 2.00 | 5816 | -0.028 | 0.004 | 0.028 | 0.000 | 1200-0.0 $\leq$ 2.00 | 5326 | -0.028 | 0.004 | 0.028 | 0.000 |
| 18 | 1170-0.0 $\leq$ 3.00 | 5516 | 0.026 | 0.004 | 0.026 | 1.000 | 1170-0.0 $\leq$ 3.00 | 4991 | 0.026 | 0.004 | 0.026 | 1.000 |
| 19 | 1528-0.0 $\leq$ 1.00 | 2705 | -0.026 | 0.004 | 0.026 | 0.000 | 1528-0.0 $\leq$ 1.00 | 2801 | -0.026 | 0.004 | 0.026 | 0.000 |
| 20 | 3.00 < 1170-0.0 $\leq$ 4.00 | 2484 | -0.026 | 0.004 | 0.026 | 0.000 | 3.00 < 1170-0.0 $\leq$ 4.00 | 3009 | -0.026 | 0.004 | 0.026 | 0.000 |

*Note.* Top-20 per side are ranked by mean absolute LIME weight ($|w|$). Values are aggregated across stratified subsamples; this table reports *rule-level* explanations, i.e., each entry in "Feature (thresholded)" is a conditional rule of the form *¡feature¿ ¡operator¿ ¡threshold¿* rather than the feature in isolation. Consequently, different thresholds of the same feature may appear as separate rows. For example, the rule *age_defined_baseline* $\leq$ *50.00* indicates that the model explanation treats individuals aged 50 or younger as a distinct subgroup with its own weight. P(+) denotes the proportion of explanations where the LIME weight was positive (i.e., contributing toward predicting the event class). Items may differ across $y = 0$ and $y = 1$ due to outcome-specific explanations.

#### 11.2 Random Forest

Table 15: LIME aggregated explanations for **Random Forest** (*per-side* Top-20 by Mean $|w|$), stratified by outcome ($y = 0$ vs. $y = 1$). The always-zero rule `6144.5` $\leq$ `1.00` was excluded.

| | Outcome = 0 (Non-event) | | | | | | Outcome = 1 (Event) | | | | | |
|---|---|---|---|---|---|---|---|---|---|---|---|---|
| **Idx** | **Feature (thresholded)** | **Count** | **Mean** | **Std** | $\lvert w\rvert$ | **P(+)** | **Feature (thresholded)** | **Count** | **Mean** | **Std** | $\lvert w\rvert$ | **P(+)** |
| 1 | age_defined_baseline ≤ 53.00 | 3126 | -0.161 | 0.011 | 0.161 | 0.000 | age_defined_baseline ≤ 53.00 | 1283 | -0.157 | 0.013 | 0.157 | 0.000 |
| 2 | age_defined_baseline > 65.00 | 1006 | 0.118 | 0.010 | 0.118 | 1.000 | age_defined_baseline > 65.00 | 2111 | 0.116 | 0.011 | 0.116 | 1.000 |
| 3 | genetic_sex ≤ 0.00 | 4574 | -0.090 | 0.009 | 0.090 | 0.000 | 0.00 < genetic_sex ≤ 1.00 | 4771 | 0.089 | 0.010 | 0.089 | 1.000 |
| 4 | 60.00 < age_defined_baseline ≤ 65.00 | 1786 | 0.090 | 0.006 | 0.090 | 1.000 | genetic_sex ≤ 0.00 | 3229 | -0.089 | 0.011 | 0.089 | 0.000 |
| 5 | 0.00 < genetic_sex ≤ 1.00 | 3426 | 0.089 | 0.010 | 0.089 | 1.000 | 60.00 < age_defined_baseline ≤ 65.00 | 2628 | 0.088 | 0.007 | 0.088 | 1.000 |
| 6 | 20161-0.0 > 12.00 | 1520 | 0.049 | 0.005 | 0.049 | 1.000 | 20161-0.0 > 12.00 | 2455 | 0.048 | 0.005 | 0.048 | 1.000 |
| 7 | 20161-0.0 ≤ 0.00 | 5597 | -0.043 | 0.004 | 0.043 | 0.000 | 20161-0.0 ≤ 0.00 | 4569 | -0.042 | 0.004 | 0.042 | 0.000 |
| 8 | 1190-0.0 ≤ 1.00 | 4665 | -0.029 | 0.005 | 0.029 | 0.000 | 1190-0.0 ≤ 1.00 | 3789 | -0.028 | 0.005 | 0.028 | 0.000 |
| 9 | 20162-0.0 ≤ 0.00 | 5409 | -0.026 | 0.004 | 0.026 | 0.000 | 20162-0.0 ≤ 0.00 | 4407 | -0.025 | 0.005 | 0.025 | 0.000 |
| 10 | 1.00 < 1190-0.0 ≤ 2.00 | 2967 | 0.024 | 0.005 | 0.024 | 1.000 | 1.00 < 1190-0.0 ≤ 2.00 | 3622 | 0.023 | 0.005 | 0.023 | 1.000 |
| 11 | 53.00 < age_defined_baseline ≤ 60.00 | 2082 | -0.022 | 0.006 | 0.022 | 0.012 | 1190-0.0 > 2.00 | 589 | 0.021 | 0.006 | 0.021 | 0.998 |
| 12 | 1190-0.0 > 2.00 | 368 | 0.022 | 0.006 | 0.022 | 1.000 | 53.00 < age_defined_baseline ≤ 60.00 | 1978 | -0.020 | 0.008 | 0.020 | 0.017 |
| 13 | 2897-0.0 > 24.00 | 1626 | 0.016 | 0.003 | 0.016 | 1.000 | 2897-0.0 > 24.00 | 2318 | 0.016 | 0.003 | 0.016 | 1.000 |
| 14 | 20162-0.0 > 0.29 | 1578 | 0.015 | 0.005 | 0.015 | 1.000 | 20162-0.0 > 0.29 | 2418 | 0.015 | 0.005 | 0.015 | 1.000 |
| 15 | 1538-0.0 ≤ 0.00 | 5024 | -0.013 | 0.005 | 0.013 | 0.001 | 2887-0.0 ≤ 0.00 | 5189 | -0.012 | 0.004 | 0.012 | 0.004 |
| 16 | 0.00 < 1538-0.0 ≤ 2.00 | 2976 | 0.013 | 0.005 | 0.013 | 1.000 | 1110-0.0 ≤ 2.00 | 3163 | 0.012 | 0.003 | 0.012 | 1.000 |
| 17 | 2887-0.0 ≤ 0.00 | 6048 | -0.012 | 0.003 | 0.012 | 0.001 | 1538-0.0 ≤ 0.00 | 4676 | -0.012 | 0.005 | 0.012 | 0.001 |
| 18 | 1110-0.0 ≤ 2.00 | 2813 | 0.012 | 0.003 | 0.012 | 1.000 | 0.00 < 1538-0.0 ≤ 2.00 | 3324 | 0.012 | 0.005 | 0.012 | 1.000 |
| 19 | 1220-0.0 ≤ 0.00 | 6151 | -0.012 | 0.003 | 0.012 | 0.000 | 1220-0.0 ≤ 0.00 | 5584 | -0.011 | 0.003 | 0.011 | 0.001 |
| 20 | 1220-0.0 > 1.00 | 209 | 0.011 | 0.006 | 0.011 | 0.962 | 2887-0.0 > 10.00 | 2108 | 0.011 | 0.006 | 0.011 | 0.999 |

*Note.* Top-20 features are ranked *separately* within $y = 0$ and $y = 1$ by the mean absolute LIME weight ($|w|$). P(+) denotes the proportion of explanations where the LIME weight was positive (i.e., contributing toward predicting the event class). Because the ranking is outcome-specific, the two sides may differ in both order and membership.

##### 11.3 XGBoost

Table 16: LIME aggregated explanations for **XGBoost** (*per-side* Top-20 by Mean |weight|), stratified by outcome ($y = 0$ vs. $y = 1$).

| | **Outcome = 0 (Non-event)** | | | | | | **Outcome = 1 (Event)** | | | | | |
|---|---|---|---|---|---|---|---|---|---|---|---|---|
| **Idx** | **Feature (thresholded)** | **Count** | **Mean** | **Std** | $\lvert w \rvert$ | **P(+)** | **Feature (thresholded)** | **Count** | **Mean** | **Std** | $\lvert w \rvert$ | **P(+)** |
| 1 | age_defined_baseline $\leq$ 53.00 | 3126 | -0.296 | 0.010 | 0.296 | 0.000 | age_defined_baseline $\leq$ 53.00 | 1283 | -0.293 | 0.013 | 0.293 | 0.000 |
| 2 | age_defined_baseline > 65.00 | 1006 | 0.242 | 0.008 | 0.242 | 1.000 | age_defined_baseline > 65.00 | 2111 | 0.240 | 0.010 | 0.240 | 1.000 |
| 3 | genetic_sex $\leq$ 0.00 | 4574 | -0.144 | 0.007 | 0.144 | 0.000 | genetic_sex $\leq$ 0.00 | 3229 | -0.143 | 0.008 | 0.143 | 0.000 |
| 4 | 0.00 < genetic_sex $\leq$ 1.00 | 3426 | 0.144 | 0.007 | 0.144 | 1.000 | 0.00 < genetic_sex $\leq$ 1.00 | 4771 | 0.143 | 0.008 | 0.143 | 1.000 |
| 5 | 60.00 < age_defined_baseline $\leq$ 65.00 | 1786 | 0.137 | 0.006 | 0.137 | 1.000 | 60.00 < age_defined_baseline $\leq$ 65.00 | 2628 | 0.137 | 0.007 | 0.137 | 1.000 |
| 6 | 1538-0.0 $\leq$ 0.00 | 5024 | -0.086 | 0.006 | 0.086 | 0.000 | 1538-0.0 $\leq$ 0.00 | 4676 | -0.086 | 0.007 | 0.086 | 0.000 |
| 7 | 0.00 < 1538-0.0 $\leq$ 2.00 | 2976 | 0.086 | 0.006 | 0.086 | 1.000 | 0.00 < 1538-0.0 $\leq$ 2.00 | 3324 | 0.085 | 0.007 | 0.085 | 1.000 |
| 8 | 20161-0.0 > 12.00 | 1520 | 0.057 | 0.008 | 0.057 | 1.000 | 20161-0.0 > 12.00 | 2455 | 0.057 | 0.009 | 0.057 | 1.000 |
| 9 | 20161-0.0 $\leq$ 0.00 | 5597 | -0.044 | 0.004 | 0.044 | 0.000 | 20161-0.0 $\leq$ 0.00 | 4569 | -0.044 | 0.005 | 0.044 | 0.000 |
| 10 | 1160-0.0 $\leq$ 6.00 | 1892 | 0.044 | 0.008 | 0.044 | 0.999 | 1160-0.0 $\leq$ 6.00 | 2209 | 0.044 | 0.008 | 0.044 | 1.000 |
| 11 | 1528-0.0 $\leq$ 1.00 | 2705 | -0.034 | 0.009 | 0.034 | 0.000 | 1528-0.0 $\leq$ 1.00 | 2801 | -0.035 | 0.010 | 0.035 | 0.000 |
| 12 | 53.00 < age_defined_baseline $\leq$ 60.00 | 2082 | -0.031 | 0.007 | 0.031 | 0.001 | 1528-0.0 > 4.00 | 1282 | 0.032 | 0.009 | 0.032 | 0.999 |
| 13 | 2.00 < 1200-0.0 $\leq$ 3.00 | 2184 | 0.031 | 0.006 | 0.031 | 1.000 | 2.00 < 1200-0.0 $\leq$ 3.00 | 2674 | 0.031 | 0.007 | 0.031 | 1.000 |
| 14 | 1528-0.0 > 4.00 | 1442 | 0.031 | 0.009 | 0.031 | 1.000 | 1200-0.0 $\leq$ 2.00 | 5326 | -0.031 | 0.006 | 0.031 | 0.000 |
| 15 | 1200-0.0 $\leq$ 2.00 | 5816 | -0.030 | 0.006 | 0.030 | 0.000 | 53.00 < age_defined_baseline $\leq$ 60.00 | 1978 | -0.029 | 0.008 | 0.029 | 0.005 |
| 16 | 1190-0.0 > 2.00 | 368 | 0.026 | 0.009 | 0.026 | 0.995 | 1190-0.0 > 2.00 | 589 | 0.025 | 0.009 | 0.025 | 0.992 |
| 17 | 2.00 < 1508-0.0 $\leq$ 3.00 | 1966 | -0.023 | 0.007 | 0.023 | 0.000 | 2.00 < 1508-0.0 $\leq$ 3.00 | 1654 | -0.022 | 0.007 | 0.022 | 0.000 |
| 18 | 1508-0.0 $\leq$ 2.00 | 5806 | 0.022 | 0.007 | 0.022 | 0.998 | 1508-0.0 $\leq$ 2.00 | 6109 | 0.022 | 0.008 | 0.022 | 0.998 |
| 19 | 6.00 < 1160-0.0 $\leq$ 7.00 | 3239 | -0.021 | 0.005 | 0.021 | 0.000 | 6.00 < 1160-0.0 $\leq$ 7.00 | 2814 | -0.021 | 0.005 | 0.021 | 0.000 |
| 20 | 1190-0.0 $\leq$ 1.00 | 4665 | -0.019 | 0.005 | 0.019 | 0.000 | 1190-0.0 $\leq$ 1.00 | 3789 | -0.020 | 0.005 | 0.020 | 0.000 |

*Note.* Top-20 features are ranked separately within $y = 0$ and $y = 1$ by the mean absolute LIME weight $|w|$. This is a *rule-level* table: each item corresponds to a conditional rule with its threshold(s) retained (e.g., "`age_defined_baseline` $\leq$ `50.00`"), aggregated over stratified subsamples. Because the ranking is outcome-specific, the two sides may differ in both order and membership. P(+) denotes the proportion of explanations where the LIME weight was positive (i.e., contributing toward predicting the event class).

### 12 Subgroup

#### 12.1 Subgroup Models Perforamnce Metrics

Complete subgroup analyses across all eight demographic strata (sex × age quartiles) are provided in Supplementary Table X, with representative results for four contrasting subgroups presented in the main text to illustrate key performance patterns.

Table 17: Subgroup performance metrics (threshold = 0.5)

| **Subgroup** | **n** | **Model** | **Prev.** | **AUC (95% CI)** | **PR-AUC** | **Acc.** | **Prec.** | **Recall** | **F1** | **Brier** |
|---|---|---|---|---|---|---|---|---|---|---|
| 0_Q1 | 14071 | LR | 0.024 | 0.658 (0.627–0.688) | 0.056 | 0.973 | 0.077 | 0.012 | 0.021 | 0.026 |
| | | RF | 0.024 | 0.617 (0.584–0.650) | 0.045 | 0.976 | 0.000 | 0.000 | 0.000 | 0.164 |
| | | XGB | 0.024 | 0.476 (0.444–0.508) | 0.023 | 0.976 | 0.000 | 0.000 | 0.000 | 0.091 |
| 0_Q2 | 12114 | LR | 0.040 | 0.630 (0.604–0.656) | 0.074 | 0.957 | 0.156 | 0.014 | 0.026 | 0.042 |
| | | RF | 0.040 | 0.609 (0.583–0.635) | 0.065 | 0.960 | 0.000 | 0.000 | 0.000 | 0.169 |
| | | XGB | 0.040 | 0.507 (0.480–0.534) | 0.043 | 0.960 | 0.000 | 0.000 | 0.000 | 0.098 |
| 0_Q3 | 14170 | LR | 0.077 | 0.600 (0.583–0.617) | 0.120 | 0.922 | 0.333 | 0.013 | 0.025 | 0.078 |
| | | RF | 0.077 | 0.585 (0.567–0.602) | 0.109 | 0.922 | 0.000 | 0.000 | 0.000 | 0.178 |
| | | XGB | 0.077 | 0.522 (0.505–0.540) | 0.084 | 0.923 | 1.000 | 0.001 | 0.002 | 0.114 |
| 0_Q4 | 10970 | LR | 0.123 | 0.571 (0.555–0.588) | 0.163 | 0.876 | 0.263 | 0.004 | 0.007 | 0.123 |
| | | RF | 0.123 | 0.553 (0.537–0.569) | 0.144 | 0.877 | 0.000 | 0.000 | 0.000 | 0.188 |
| | | XGB | 0.123 | 0.514 (0.497–0.530) | 0.131 | 0.877 | 0.200 | 0.001 | 0.001 | 0.134 |
| 1_Q1 | 11526 | LR | 0.045 | 0.608 (0.583–0.633) | 0.074 | 0.953 | 0.158 | 0.012 | 0.022 | 0.047 |
| | | RF | 0.045 | 0.579 (0.554–0.605) | 0.067 | 0.878 | 0.080 | 0.162 | 0.107 | 0.212 |
| | | XGB | 0.045 | 0.525 (0.500–0.551) | 0.050 | 0.920 | 0.055 | 0.048 | 0.051 | 0.159 |
| 1_Q2 | 9213 | LR | 0.089 | 0.584 (0.563–0.603) | 0.119 | 0.909 | 0.206 | 0.009 | 0.016 | 0.090 |
| | | RF | 0.089 | 0.561 (0.540–0.581) | 0.113 | 0.829 | 0.136 | 0.172 | 0.152 | 0.219 |
| | | XGB | 0.089 | 0.508 (0.488–0.530) | 0.089 | 0.880 | 0.085 | 0.036 | 0.050 | 0.168 |
| 1_Q3 | 10511 | LR | 0.151 | 0.583 (0.569–0.599) | 0.204 | 0.848 | 0.356 | 0.010 | 0.020 | 0.150 |
| | | RF | 0.151 | 0.574 (0.559–0.590) | 0.194 | 0.780 | 0.239 | 0.209 | 0.223 | 0.226 |
| | | XGB | 0.151 | 0.528 (0.513–0.544) | 0.165 | 0.829 | 0.192 | 0.042 | 0.069 | 0.180 |
| 1_Q4 | 9254 | LR | 0.204 | 0.569 (0.555–0.584) | 0.248 | 0.795 | 0.290 | 0.005 | 0.009 | 0.202 |
| | | RF | 0.204 | 0.562 (0.547–0.577) | 0.245 | 0.727 | 0.275 | 0.206 | 0.235 | 0.231 |
| | | XGB | 0.204 | 0.509 (0.494–0.524) | 0.214 | 0.780 | 0.251 | 0.039 | 0.067 | 0.193 |

#### 12.2 Top 20 SHAP feature in Age = Q1, Sex = 0

Table 18: Subgroup SHAP Top-20 Features (Age = Q1, Sex = 0). Scores are mean absolute SHAP values.

| Rank | LR | RF | XGB |
|---|---|---|---|
| 1 | Age at baseline (age_defined_baseline) | Age at baseline (age_defined_baseline) | Age at baseline (age_defined_baseline) |
| 2 | Sex (genetic_sex) | Sex (genetic_sex) | Sleep duration (1160) |
| 3 | Time from waking to first cigarette (3466) | Length of mobile phone use (1110) | Sex (genetic_sex) |
| 4 | Exposure to tobacco smoke at home (1269) | Nap during day (1190) | Major dietary changes in the last 5 years (1538) |
| 5 | Smoking/smokers in household (1259) | Sleep duration (1160) | Getting up in morning (1170) |
| 6 | Major dietary changes in the last 5 years (1538) | Pack years adult smoking as proportion of lifespan (20162) | Summed days of activity (22033) |
| 7 | Nap during day (1190) | Pack years of smoking (20161) | Cheese intake (1408) |
| 8 | Getting up in morning (1170) | Summed MET minutes/week for all activity (22040) | Hot drink temperature (1518) |
| 9 | Insomnia / sleeplessness (1200) | Summed minutes activity (22034) | Summed minutes activity (22034) |
| 10 | Cheese intake (1408) | Insomnia / sleeplessness (1200) | Insomnia / sleeplessness (1200) |
| 11 | Ever smoked (20160) | Major dietary changes in the last 5 years (1538) | Water intake (1528) |
| 12 | Never eat eggs, dairy, wheat, sugar (6144.5) | Cheese intake (1408) | Never eat eggs, dairy, wheat, sugar (6144.5) |
| 13 | Pack years of smoking (20161) | Snoring (1210) | Vigorous MET minutes/week (22039) |
| 14 | Water intake (1528) | Bread intake (1438) | Passive smoke exposure outside home (1279) |
| 15 | Age started smoking in former smokers (2867) | Daytime dozing / sleeping (narcolepsy) (1220) | Pack years of smoking (20161) |
| 16 | Pack years adult smoking as proportion of lifespan (20162) | Vigorous MET minutes/week (22039) | Nap during day (1190) |
| 17 | Bread type (1448) | Past tobacco smoking (1249) | Oily fish intake (1329) |
| 18 | Weekly usage of mobile phone in last 3 months (1120) | Never eat eggs, dairy, wheat, sugar (6144.5) | Bread type (1448) |
| 19 | Type of tobacco previously smoked (2877) | Salad / raw vegetable intake (1299) | Salad / raw vegetable intake (1299) |
| 20 | Summed days of activity (22033) | Processed meat intake (1349) | Snoring (1210) |

*Note.* Rankings are based on mean absolute SHAP values within subgroup **Age = Q1, Sex = 0**. Features are mapped to UK Biobank variable names, with option-level codes (e.g., 6144.5 for diet, 3456 for cigarettes/day) retained.

#### 12.3 Top 20 SHAP feature in Age = Q2, Sex = 0

Table 19: Subgroup SHAP Top-20 Features (Age = Q2, Sex = 0). Scores are mean absolute SHAP values.

| Rank | LR | RF | XGB |
|---|---|---|---|
| 1 | Sex (genetic_sex) | Age at baseline (age_defined_baseline) | Age at baseline (age_defined_baseline) |
| 2 | Age at baseline (age_defined_baseline) | Sex (genetic_sex) | Sleep duration (1160) |
| 3 | Time from waking to first cigarette (3466) | Length of mobile phone use (1110) | Sex (genetic_sex) |
| 4 | Exposure to tobacco smoke at home (1269) | Nap during day (1190) | Major dietary changes in last 5 years (1538) |
| 5 | Smoking/smokers in household (1259) | Sleep duration (1160) | Getting up in morning (1170) |
| 6 | Major dietary changes in the last 5 years (1538) | Pack years as proportion of lifespan (20162) | Summed days of activity (22033) |
| 7 | Why stopped smoking (6157; code -888) | Pack years of smoking (20161) | Cheese intake (1408) |
| 8 | Nap during day (1190) | Summed MET minutes per week for all activity (22040) | Hot drink temperature (1518) |
| 9 | Never eat eggs, dairy, wheat, sugar (6144.5) | Summed minutes activity (22034) | Summed minutes activity (22034) |
| 10 | Pack years of smoking (20161) | Insomnia / sleeplessness (1200) | Insomnia / sleeplessness (1200) |
| 11 | Sleeplessness / insomnia (1200) | Major dietary changes in the last 5 years (1538) | Water intake (1528) |
| 12 | Cheese intake (1408) | Cheese intake (1408) | Never eat eggs, dairy, wheat, sugar (6144.5) |
| 13 | Getting up in morning (1170) | Snoring (1210) | Vigorous MET minutes/week (22039) |
| 14 | Ever smoked (20160) | Bread intake (1438) | Exposure to tobacco smoke outside home (1279) |
| 15 | Age started smoking in former smokers (2867) | Daytime dozing / sleeping (narcolepsy) (1220) | Pack years of smoking (20161) |
| 16 | Water intake (1528) | Summed days activity (22033) | Nap during day (1190) |
| 17 | Pack years as proportion of lifespan (20162) | MET minutes per week for vigorous activity (22039) | Oily fish intake (1329) |
| 18 | Bread type (1448) | Past tobacco smoking (1249) | Bread type (1448) |
| 19 | Type of tobacco previously smoked (2877) | Never eat eggs, dairy, wheat, sugar (6144.5) | Salad / raw vegetable intake (1299) |
| 20 | Weekly usage of mobile phone in last 3 months (1120) | Salad / raw vegetable intake (1299) | Snoring (1210) |

*Note.* Rankings are based on mean absolute SHAP values within subgroup **Age = Q2, Sex = 0**. Features are mapped to UK Biobank variable names, with option-level codes (e.g., 6144.5 for diet, 3456 for cigarettes/day) retained.

#### 12.4 Top 20 SHAP feature in Age = Q3, Sex = 0

Table 20: Subgroup SHAP Top-20 Features (Age = Q3, Sex = 0). Scores are mean absolute SHAP values.

| Rank | LR | RF | XGB |
|---|---|---|---|
| 1 | Age at baseline (age_defined_baseline) | Age at baseline (age_defined_baseline) | Age at baseline (age_defined_baseline) |
| 2 | Sex (genetic_sex) | Sex (genetic_sex) | Sleep duration (1160) |
| 3 | Time from waking to first cigarette (3466) | Length of mobile phone use (1110) | Sex (genetic_sex) |
| 4 | Major dietary changes in the last 5 years (1538) | Nap during day (1190) | Major dietary changes in the last 5 years (1538) |
| 5 | Exposure to tobacco smoke at home (1269) | Sleep duration (1160) | Getting up in morning (1170) |
| 6 | Nap during day (1190) | Pack years adult smoking as proportion of lifespan (20162) | Summed days of activity (22033) |
| 7 | Never eat eggs, dairy, wheat, sugar (6144.5) | Pack years of smoking (20161) | Cheese intake (1408) |
| 8 | Smoking/smokers in household (1259) | Summed MET minutes/week for all activity (22040) | Hot drink temperature (1518) |
| 9 | Pack years of smoking (20161) | Summed minutes activity (22034) | Summed minutes activity (22034) |
| 10 | Age started smoking in former smokers (2867) | Insomnia / sleeplessness (1200) | Insomnia / sleeplessness (1200) |
| 11 | Cheese intake (1408) | Major dietary changes in the last 5 years (1538) | Water intake (1528) |
| 12 | Ever smoked (20160) | Cheese intake (1408) | Never eat eggs, dairy, wheat, sugar (6144.5) |
| 13 | Insomnia / sleeplessness (1200) | Bread intake (1438) | Vigorous MET minutes/week (22039) |
| 14 | Getting up in morning (1170) | Snoring (1210) | Passive smoke exposure outside home (1279) |
| 15 | Pack years adult smoking as proportion of lifespan (20162) | Daytime dozing / sleeping (narcolepsy) (1220) | Pack years of smoking (20161) |
| 16 | Water intake (1528) | Never eat eggs, dairy, wheat, sugar (6144.5) | Nap during day (1190) |
| 17 | Type of tobacco previously smoked (2877) | Summed days of activity (22033) | Oily fish intake (1329) |
| 18 | Bread type (1448) | Vigorous MET minutes/week (22039) | Bread type (1448) |
| 19 | Weekly usage of mobile phone in last 3 months (1120) | Past tobacco smoking (1249) | Salad / raw vegetable intake (1299) |
| 20 | Summed days of activity (22033) | Age stopped smoking (2897) | Snoring (1210) |

*Note.* Rankings are based on mean absolute SHAP values within subgroup **Age = Q3, Sex = 0**. Features are mapped to UK Biobank variable names, with option-level codes (e.g., 6144.5 for diet, 3456 for cigarettes/day) retained.

##### 12.5 Top 20 SHAP feature in Age = Q4, Sex = 0

Table 21: Subgroup SHAP Top-20 Features (Age = Q4, Sex = 0). Scores are mean absolute SHAP values.

| Rank | LR | RF | XGB |
|---|---|---|---|
| 1 | Age at baseline (age_defined_baseline) | Age at baseline (age_defined_baseline) | Age at baseline (age_defined_baseline) |
| 2 | Sex (genetic_sex) | Sex (genetic_sex) | Sleep duration (1160) |
| 3 | Time from waking to first cigarette (3466) | Length of mobile phone use (1110) | Sex (genetic_sex) |
| 4 | Never eat eggs, dairy, wheat, sugar (6144.5) | Nap during day (1190) | Major dietary changes in the last 5 years (1538) |
| 5 | Nap during day (1190) | Sleep duration (1160) | Getting up in morning (1170) |
| 6 | Major dietary changes in the last 5 years (1538) | Pack years adult smoking as proportion of lifespan (20162) | Summed days of activity (22033) |
| 7 | Pack years of smoking (20161) | Pack years of smoking (20161) | Cheese intake (1408) |
| 8 | Exposure to tobacco smoke at home (1269) | Summed MET minutes/week for all activity (22040) | Hot drink temperature (1518) |
| 9 | Smoking/smokers in household (1259) | Summed minutes activity (22034) | Summed minutes activity (22034) |
| 10 | Age started smoking in former smokers (2867) | Insomnia / sleeplessness (1200) | Insomnia / sleeplessness (1200) |
| 11 | Cheese intake (1408) | Daytime dozing / sleeping (narcolepsy) (1220) | Water intake (1528) |
| 12 | Ever smoked (20160) | Cheese intake (1408) | Never eat eggs, dairy, wheat, sugar (6144.5) |
| 13 | Getting up in morning (1170) | Bread intake (1438) | Vigorous MET minutes/week (22039) |
| 14 | Insomnia / sleeplessness (1200) | Snoring (1210) | Passive smoke exposure outside home (1279) |
| 15 | Pack years adult smoking as proportion of lifespan (20162) | Never eat eggs, dairy, wheat, sugar (6144.5) | Pack years of smoking (20161) |
| 16 | Type of tobacco previously smoked (2877) | Major dietary changes in the last 5 years (1538) | Nap during day (1190) |
| 17 | Bread type (1448) | Summed days of activity (22033) | Oily fish intake (1329) |
| 18 | Water intake (1528) | Vigorous MET minutes/week (22039) | Salad / raw vegetable intake (1299) |
| 19 | Weekly usage of mobile phone in last 3 months (1120) | Past tobacco smoking (1249) | Snoring (1210) |
| 20 | Summed days of activity (22033) | Salad / raw vegetable intake (1299) | Bread type (1448) |

*Note.* Rankings are based on mean absolute SHAP values within subgroup **Age = Q4, Sex = 0**. Features are mapped to UK Biobank variable names, with option-level codes (e.g., 6144.5 for diet, 3456 for cigarettes/day) retained.

##### 12.6 Top 20 SHAP feature in Age = Q1, Sex = 1

Table 22: Top-20 SHAP features for subgroup **Age = Q1, Sex = 1**, across three models (Logistic Regression, Random Forest, XGBoost).

| Rank | Logistic Regression (SHAP) | Random Forest (SHAP) | XGBoost (SHAP) |
|---|---|---|---|
| 1 | Age at baseline (age_defined_baseline) | Age at baseline (age_defined_baseline) | Age at baseline (age_defined_baseline) |
| 2 | Sex (genetic_sex) | Sex (genetic_sex) | Sex (genetic_sex) |
| 3 | Time from waking to first cigarette (3466) | Nap during day (1190) | Sleep duration (1160) |
| 4 | Exposure to tobacco smoke at home (1269) | Length of mobile phone use (1110) | Major dietary changes in the last 5 years (1538) |
| 5 | Smoking/smokers in household (1259) | Pack years adult smoking as proportion of lifespan (20162) | Getting up in morning (1170) |
| 6 | Major dietary changes in the last 5 years (1538) | Pack years of smoking (20161) | Summed days activity (22033) |
| 7 | Nap during day (1190) | Summed minutes activity (22034) | Cheese intake (1408) |
| 8 | Pack years of smoking (20161) | Sleep duration (1160) | Hot drink temperature (1518) |
| 9 | Insomnia / sleeplessness (1200) | Snoring (1210) | Summed minutes activity (22034) |
| 10 | Pack years adult smoking as proportion of lifespan (20162) | Summed MET minutes/week for all activity (22040) | Water intake (1528) |
| 11 | Weekly usage of mobile phone in last 3 months (1120) | Bread intake (1438) | Insomnia / sleeplessness (1200) |
| 12 | Ever smoked (20160) | Past tobacco smoking (1249) | Major dietary changes in the last 5 years (1538) |
| 13 | Cheese intake (1408) | Major dietary changes in the last 5 years (1538) | Getting up in morning (1170) |
| 14 | Getting up in morning (1170) | Cheese intake (1408) | Never eat eggs, dairy, wheat, sugar (6144.5) |
| 15 | Age started smoking in former smokers (2867) | Insomnia / sleeplessness (1200) | Vigorous MET minutes/week (22039) |
| 16 | Never eat eggs, dairy, wheat, sugar (6144.5) | Salad / raw vegetable intake (1299) | Passive smoke exposure outside home (1279) |
| 17 | Water intake (1528) | Weekly usage of mobile phone in last 3 months (1120) | Pack years of smoking (20161) |
| 18 | Bread type (1448) | Age stopped smoking (2897) | Nap during day (1190) |
| 19 | Summed days activity (22033) | Daytime dozing / sleeping (narcolepsy) (1220) | Oily fish intake (1329) |
| 20 | Type of tobacco previously smoked (2877) | Vigorous MET minutes/week (22039) | Snoring (1210) |

*Note.* Rankings are based on mean absolute SHAP values within subgroup **Age = Q1, Sex = 1**. Features are mapped to UK Biobank variable names, retaining option-level codes (e.g., 6144.5 for diet, 2897 for smoking cessation).

#### 12.7 Top 20 SHAP feature in Age = Q2, Sex = 1

Table 23: Top-20 SHAP features for subgroup **Age = Q2, Sex = 1**, across three models (Logistic Regression, Random Forest, XGBoost).

| Rank | Logistic Regression (SHAP) | Random Forest (SHAP) | XGBoost (SHAP) |
|---|---|---|---|
| 1 | Sex (genetic_sex) | Age at baseline (age_defined_baseline) | Age at baseline (age_defined_baseline) |
| 2 | Age at baseline (age_defined_baseline) | Sex (genetic_sex) | Sex (genetic_sex) |
| 3 | Time from waking to first cigarette (3466) | Nap during day (1190) | Sleep duration (1160) |
| 4 | Exposure to tobacco smoke at home (1269) | Length of mobile phone use (1110) | Major dietary changes in the last 5 years (1538) |
| 5 | Smoking/smokers in household (1259) | Pack years adult smoking as proportion of lifespan (20162) | Getting up in morning (1170) |
| 6 | Nap during day (1190) | Pack years of smoking (20161) | Summed days activity (22033) |
| 7 | Pack years of smoking (20161) | Summed minutes activity (22034) | Cheese intake (1408) |
| 8 | Major dietary changes in the last 5 years (1538) | Sleep duration (1160) | Hot drink temperature (1518) |
| 9 | Never eat eggs, dairy, wheat, sugar (6144.5) | Snoring (1210) | Summed minutes activity (22034) |
| 10 | Insomnia / sleeplessness (1200) | Summed MET minutes/week for all activity (22040) | Water intake (1528) |
| 11 | Age started smoking in former smokers (2867) | Bread intake (1438) | Insomnia / sleeplessness (1200) |
| 12 | Pack years adult smoking as proportion of lifespan (20162) | Past tobacco smoking (1249) | Never eat eggs, dairy, wheat, sugar (6144.5) |
| 13 | Ever smoked (20160) | Major dietary changes in the last 5 years (1538) | Vigorous MET minutes/week (22039) |
| 14 | Cheese intake (1408) | Weekly usage of mobile phone in last 3 months (1120) | Passive smoke exposure outside home (1279) |
| 15 | Getting up in morning (1170) | Insomnia / sleeplessness (1200) | Pack years of smoking (20161) |
| 16 | Water intake (1528) | Daytime dozing / sleeping (narcolepsy) (1220) | Nap during day (1190) |
| 17 | Number of cigarettes currently smoked daily (3456) | Salad / raw vegetable intake (1299) | Oily fish intake (1329) |
| 18 | Bread type (1448) | Age stopped smoking (2897) | Salad / raw vegetable intake (1299) |
| 19 | Weekly usage of mobile phone in last 3 months (1120) | Never eat eggs, dairy, wheat, sugar (6144.5) | Snoring (1210) |
| 20 | Type of tobacco previously smoked (2877) | Vigorous MET minutes/week (22039) | Bread type (1448) |

*Note.* Rankings are based on mean absolute SHAP values within subgroup **Age = Q2, Sex = 1**. Features are mapped to UK Biobank variable names, retaining option-level codes (e.g., 6144.5 for diet, 3456 for cigarettes/day).

#### 12.8 Top 20 SHAP feature in Age = Q3, Sex = 1

Table 24: Top-20 SHAP features for subgroup **Age = Q3, Sex = 1**, across three models (Logistic Regression, Random Forest, XGBoost).

| Rank | Logistic Regression (SHAP) | Random Forest (SHAP) | XGBoost (SHAP) |
|---|---|---|---|
| 1 | Age at baseline (age_defined_baseline) | Age at baseline (age_defined_baseline) | Age at baseline (age_defined_baseline) |
| 2 | Sex (genetic_sex) | Sex (genetic_sex) | Sex (genetic_sex) |
| 3 | Time from waking to first cigarette (3466) | Nap during day (1190) | Sleep duration (1160) |
| 4 | Exposure to tobacco smoke at home (1269) | Length of mobile phone use (1110) | Major dietary changes in the last 5 years (1538) |
| 5 | Pack years of smoking (20161) | Pack years adult smoking as proportion of lifespan (20162) | Getting up in morning (1170) |
| 6 | Smoking/smokers in household (1259) | Pack years of smoking (20161) | Summed days activity (22033) |
| 7 | Nap during day (1190) | Summed minutes activity (22034) | Cheese intake (1408) |
| 8 | Major dietary changes in the last 5 years (1538) | Sleep duration (1160) | Hot drink temperature (1518) |
| 9 | Never eat eggs, dairy, wheat, sugar (6144.5) | Summed MET minutes/week for all activity (22040) | Summed minutes activity (22034) |
| 10 | Age started smoking in former smokers (2867) | Bread intake (1438) | Water intake (1528) |
| 11 | Insomnia / sleeplessness (1200) | Snoring (1210) | Insomnia / sleeplessness (1200) |
| 12 | Pack years adult smoking as proportion of lifespan (20162) | Past tobacco smoking (1249) | Never eat eggs, dairy, wheat, sugar (6144.5) |
| 13 | Number of cigarettes currently smoked daily (3456) | Weekly usage of mobile phone in last 3 months (1120) | Vigorous MET minutes/week (22039) |
| 14 | Ever smoked (20160) | Cheese intake (1408) | Passive smoke exposure outside home (1279) |
| 15 | Cheese intake (1408) | Major dietary changes in the last 5 years (1538) | Pack years of smoking (20161) |
| 16 | Type of tobacco previously smoked (2877) | Daytime dozing / sleeping (narcolepsy) (1220) | Nap during day (1190) |
| 17 | Getting up in morning (1170) | Insomnia / sleeplessness (1200) | Oily fish intake (1329) |
| 18 | Water intake (1528) | Never eat eggs, dairy, wheat, sugar (6144.5) | Salad / raw vegetable intake (1299) |
| 19 | Bread type (1448) | Salad / raw vegetable intake (1299) | Snoring (1210) |
| 20 | Weekly usage of mobile phone in last 3 months (1120) | Age stopped smoking (2897) | Bread type (1448) |

*Note.* Rankings are based on mean absolute SHAP values within subgroup **Age = Q3, Sex = 1**. Features are mapped to UK Biobank variable names, with option-level codes (e.g., 6144.5 for diet, 3456 for cigarettes/day) retained.

#### 12.9 Top 20 SHAP feature in Age = Q4, Sex = 1

Table 25: Top-20 SHAP features for subgroup **Age = Q4, Sex = 1**, across three models (Logistic Regression, Random Forest, XGBoost).

| Rank | Logistic Regression (SHAP) | Random Forest (SHAP) | XGBoost (SHAP) |
|---|---|---|---|
| 1 | Age at baseline (age_defined_baseline) | Age at baseline (age_defined_baseline) | Age at baseline (age_defined_baseline) |
| 2 | Sex (genetic_sex) | Sex (genetic_sex) | Sex (genetic_sex) |
| 3 | Pack years of smoking (20161) | Nap during day (1190) | Sleep duration (1160) |
| 4 | Nap during day (1190) | Length of mobile phone use (1110) | Major dietary changes in the last 5 years (1538) |
| 5 | Time from waking to first cigarette (3466) | Pack years adult smoking as proportion of lifespan (20162) | Getting up in morning (1170) |
| 6 | Exposure to tobacco smoke at home (1269) | Pack years of smoking (20161) | Summed days activity (22033) |
| 7 | Age started smoking in former smokers (2867) | Summed minutes activity (22034) | Cheese intake (1408) |
| 8 | Smoking/smokers in household (1259) | Sleep duration (1160) | Hot drink temperature (1518) |
| 9 | Never eat eggs, dairy, wheat, sugar (6144.5) | Summed MET minutes/week for all activity (22040) | Water intake (1528) |
| 10 | Major dietary changes in the last 5 years (1538) | Bread intake (1438) | Summed minutes activity (22034) |
| 11 | Number of cigarettes currently smoked daily (3456) | Snoring (1210) | Insomnia / sleeplessness (1200) |
| 12 | Insomnia / sleeplessness (1200) | Past tobacco smoking (1249) | Never eat eggs, dairy, wheat, sugar (6144.5) |
| 13 | Type of tobacco previously smoked (2877) | Weekly usage of mobile phone in last 3 months (1120) | Vigorous MET minutes/week (22039) |
| 14 | Pack years adult smoking as proportion of lifespan (20162) | Daytime dozing / sleeping (narcolepsy) (1220) | Passive smoke exposure outside home (1279) |
| 15 | Getting up in morning (1170) | Insomnia / sleeplessness (1200) | Pack years of smoking (20161) |
| 16 | Ever smoked (20160) | Cheese intake (1408) | Nap during day (1190) |
| 17 | Cheese intake (1408) | Never eat eggs, dairy, wheat, sugar (6144.5) | Oily fish intake (1329) |
| 18 | Water intake (1528) | Major dietary changes in the last 5 years (1538) | Snoring (1210) |
| 19 | Bread type (1448) | Salad / raw vegetable intake (1299) | Salad / raw vegetable intake (1299) |
| 20 | Weekly usage of mobile phone in last 3 months (1120) | Age stopped smoking (2897) | Bread type (1448) |

*Note.* Rankings are based on mean absolute SHAP values within subgroup **Age = Q4, Sex = 1**. Features are mapped to UK Biobank variable names, with option-level codes (e.g., 6144.5 for diet, 3456 for cigarettes/day) retained.
